# Demographic Calibration Gaps in Breast Cancer Risk Prediction: Introducing the Demographic Calibration Gap Score

**DOI:** 10.64898/2026.06.17.26355900

**Authors:** Michael O. Eniolade

## Abstract

**Background:** Most breast cancer prediction studies skip calibration reporting entirely. Fewer still examine calibration by demographic subgroup. Predicted probabilities that are systematically off for specific racial or gender groups produce biased clinical decisions, and aggregate statistics will not catch that.

**Objective:** To introduce the Demographic Calibration Gap Score (DCGS), a metric that measures how much calibration error varies across demographic subgroups, and to show how it performs across five classifiers, four calibration conditions, and two datasets.

**Methods:** Five classifiers were trained on the Wisconsin Diagnostic Breast Cancer dataset (n=569) and evaluated on a breast cancer cohort from MIMIC-IV (n=1,316). Three global calibration methods were applied: no calibration, Platt scaling, and isotonic regression. A fourth condition, subgroup-targeted Platt scaling, was applied to the MIMIC cohort. DCGS was computed as DCGS_range_ = max*_k_*(ECE*_k_*) *−* min*_k_*(ECE*_k_*) across racial and gender subgroups, with 95% bootstrap confidence intervals. Conformal prediction coverage and Demographic Coverage Gap (DCG) were reported.

**Results:** On Wisconsin, all five models achieved AUROC above 0.98 and ECE below 0.12. Performance fell sharply on the MIMIC external cohort: AUROC dropped to 0.45-0.57 for base and globally calibrated variants, confirming distributional shift. DCGS exceeded the 0.05 clinical significance threshold in 28 of 40 model-calibration combinations on the race axis. Neither global Platt nor isotonic calibration reliably reduced DCGS below that threshold. Conformal coverage collapsed to roughly 25% on MIMIC, and racial DCG exceeded 0.15 for all 20 model-variant combinations.

**Conclusions:** Reducing population-level ECE through global recalibration does not reliably close demographic calibration gaps. DCGS gives researchers a direct, standardized way to detect and report those disparities. Code and the DCGS computation library are released as open-source Python under the MIT License.

## 1 Introduction

Breast cancer is one of the most frequently diagnosed cancers in the world. Roughly 300,000 new cases are expected in the United States each year. Over the past decade, machine learning models trained on cytology images, genomic data, and clinical variables have come with impressive discrimination scores attached [31–34]. AUROC curves look good in paper after paper.

Calibration is the part that gets left out. A calibrated model assigns a 70% probability of malignancy only when roughly 70% of similar patients are truly malignant. Without that alignment, a clinician reading the probability is working with a number that sounds precise but does not mean what it appears to mean.

A systematic review of machine learning approaches in breast cancer prediction found that only 3.2% of studies assessed calibration at all [1]. A separate review rated more than half of 107 prediction models as high risk of bias, with insufficient calibration reporting as a primary reason [2]. A further systematic review of ML prediction model studies found widespread gaps in methodological conduct and reporting [35]. Among studies that do report calibration, the reporting is almost always at the population level. A global ECE of 0.04 says the model is reasonably well calibrated on average. Nothing about that number tells you whether the error is spread evenly across racial and gender groups.

The uneven distribution carries real stakes. Black women in the United States face higher breast cancer mortality than White women despite lower incidence. The disparity holds after controlling for stage, treatment access, and tumor biology [3, 4]. A risk model calibrated poorly for Black patients assigns systematically biased probabilities. Patients end up below clinical decision thresholds for the wrong reasons, and that harm does not show up in the aggregate ECE.

No published study in breast cancer AI has named a metric specifically for demographic calibration disparity. Fairness metrics like equalized odds address whether a model ranks patients equally across groups [5]. Calibration equity is a different question. A model with perfect equalized odds still assigns probabilities 15 percentage points too high for one racial group and 15 points too low for another [6].

We introduce the Demographic Calibration Gap Score (DCGS). DCGS measures how much ECE varies across demographic subgroups, defined as max*_k_*(ECE*_k_*) *−* min*_k_*(ECE*_k_*) across K qualifying subgroups. A spread version uses standard deviation for sensitivity to intermediate gaps. A DCGS above 0.05 is treated as clinically meaningful.

DCGS is tested on five classifiers trained on the Wisconsin Diagnostic Breast Cancer dataset and evaluated on an emergency department cohort from MIMIC-IV [7, 8]. The two-dataset design is a deliberate stress test. Wisconsin features are FNA nucleus measurements. MIMIC features are triage vitals. Models validated internally get applied to a population with known demographic composition and a completely different feature structure.

We also extend the conformal prediction framework from prior work [9], computing a Demographic Coverage Gap (DCG) to measure how unequally prediction set coverage distributes across racial subgroups under the same threshold. All code, including the DCGS computation library, is released as open-source under the MIT License.

## 2 Background

### Calibration in clinical AI

Calibration is the alignment between predicted probabilities and observed event rates. A well-calibrated model produces estimates usable directly as decision inputs. The standard tool for assessing this is a reliability diagram, which plots mean predicted probability against the observed fraction of positives within bins. The Expected Calibration Error (ECE) summarizes the diagram numerically [10, 11, 28, 29].

Two standard post-hoc approaches exist for correcting miscalibration. Platt scaling fits a logistic regression on the base model’s output scores using held-out validation data [12]. Isotonic regression fits a monotone piecewise-constant function [13]. Both are applied globally, fitting one calibration function across the entire population.

Global calibration works when the population is homogeneous. When subgroups have different feature distributions or base rates, a global calibrator reduces aggregate ECE while potentially widening subgroup gaps. DCGS is built to detect exactly that.

### Fairness and calibration equity

The AI fairness literature mostly focuses on whether predictive performance is equal across groups. Hardt et al. formalized equalized odds [5]. Multicalibration, from Hebert-Johnson et al., requires good calibration within every auditable subgroup simultaneously [6]. La Cava et al. built proportional multicalibration as a scalable variant [14]. Shui et al. examined calibration bias in medical imaging [15]. Chen et al. reviewed algorithmic fairness in medicine broadly [16].

DCGS takes a simpler path than multicalibration. Computing ECE per subgroup and summarizing the spread requires standard calibration tools and subgroup labels. The approach fits into any existing evaluation pipeline. Reporting it adds one row to a calibration table.

### Disparities in breast cancer outcomes

Racial disparities in breast cancer mortality are well documented. Black women are approximately 40% more likely to die from breast cancer than White women despite lower incidence, a gap that persists after controlling for stage, treatment, and tumor biology [3, 17]. Yedjou et al. identified structural and biological contributors [17]. Hines et al. examined insurance type as a socioeconomic proxy [18].

In breast cancer AI research, Soltan and Washington found that deep learning models for breast cancer stage classification consistently perform better for White patients than for non-White patients, and post-processing fairness corrections reduce but do not eliminate that gap [19]. Parikh et al. developed machine learning models to predict six-month mortality across a large cancer population, establishing a template for high-stakes clinical risk scoring that subsequent work has examined for racial equity [20]. Dankwa-Mullan and Weeraratne catalogued AI-driven disparities in cancer care broadly [21]. Obermeyer et al. showed how miscalibrated algorithms propagate existing health disparities in a general clinical setting [22].

### Conformal prediction and coverage equity

Conformal prediction produces prediction sets with marginal coverage guarantees that are distribution-free [23]. In the split conformal framework, nonconformity scores from a held-out calibration set define a threshold ensuring the true label falls within the prediction set at a target rate on new data. The marginal guarantee holds over the full population but does not extend to subgroups automatically.

Zhou and Sesia examined conformal prediction with equalized coverage requirements, showing that subgroup-specific calibration is needed when coverage equity matters [24]. Lu et al. studied equalized coverage in medical imaging specifically [25]. The Demographic Coverage Gap metric follows that framework directly.

## 3 Methods

### Study design

The design uses two datasets for external validation. Five classifiers are trained and calibrated on the Wisconsin Diagnostic Breast Cancer dataset, a publicly available FNA cytology dataset with no demographic variables. External validation uses a breast cancer cohort from MIMIC-IV that provides race and gender for every patient. The feature mismatch between datasets is deliberate. It reflects realistic deployment: a model trained in one clinical context, then applied to patients from a different population.

### Primary dataset: Wisconsin Diagnostic Breast Cancer

The Wisconsin Diagnostic Breast Cancer dataset (UCI repository, id=17, n=569) contains 30 numeric features computed from fine needle aspirate nucleus images: radius, texture, perimeter, area, smoothness, compactness, concavity, symmetry, and fractal dimension, each as mean, standard error, and worst-case value [26]. Malignant cases (n=212) were labeled 1 and benign cases (n=357) were labeled 0. The dataset was split into training (60%, n=341), validation (20%, n=114), and test (20%, n=114) sets using stratified random sampling with random_state=42. A StandardScaler was fit on the training set only and applied to all splits. Training-set feature medians were saved for MIMIC feature imputation.

### External validation cohort: MIMIC-IV

MIMIC-IV is a large de-identified clinical database from Beth Israel Deaconess Medical Center in Boston [7, 8]. The analysis uses the MIMIC-IV emergency department module (version 2.2). Breast cancer patients were identified by ICD-10 code C50.x and ICD-9 codes 174.x and 175.x. All stays without any malignancy or neoplasm code were eligible as controls and were randomly sampled at a 3:1 ratio to cases, yielding a final cohort of 1,316 patients (329 positive, 987 negative).

Race was standardized to five groups: WHITE, BLACK, HISPANIC, ASIAN, and OTHER. The distribution was: WHITE 55.0%, BLACK 22.9%, OTHER 8.8%, HISPANIC 8.7%, ASIAN 4.6%. Gender was encoded as binary (F/M) from the ED stays table.

Feature harmonisation mapped nine MIMIC-IV triage variables to the first nine Wisconsin feature positions: heart rate to position 0 (mean radius), temperature to position 1 (mean texture), systolic BP to position 2 (mean perimeter), diastolic BP to position 3 (mean area), oxygen saturation to position 4 (mean smoothness), respiratory rate to position 5 (mean compactness), pain score to position 6 (mean concavity), acuity level to position 7 (mean concave points), and gender encoding to position 8 (mean symmetry). Non-numeric pain values were treated as missing and replaced with Wisconsin training-set medians. The remaining 21 Wisconsin positions were filled with training-set medians, and the full 30-feature vector was scaled with the saved Wisconsin StandardScaler.

For evaluating subgroup-targeted Platt scaling, the MIMIC cohort was split 50/50 into a calibration half (n=658) and an evaluation half (n=658), stratified by label.

### Model training and calibration

Five classifiers were trained using GridSearchCV with five-fold stratified cross-validation on the Wisconsin training set, optimizing AUROC: logistic regression (LR), random forest (RF), support vector machine (SVM, probability=True), Gaussian Naive Bayes (NB), and XGBoost (XGB). Best hyperparameters for each model are available in the code repository at github.com/MichaelEnny/breast-cancer-calibration-gaps.

Three calibration conditions were applied. Base: no post-hoc calibration. Platt: sigmoid calibration fitted on the Wisconsin validation set using the pre-fitted base model (sklearn CalibratedClassifierCV, cv=None, ensemble=False). Isotonic: isotonic regression fitted on the same validation set. A fourth condition, subgroup-targeted Platt (subgroup_platt), was applied only to the MIMIC evaluation half. It fits separate Platt scalers per racial group using MIMIC calibration-half data, falling back to the global Platt scaler for groups with fewer than 30 calibration instances.

### The DCGS metric

DCGS is computed in two steps. For each qualifying demographic subgroup (minimum n=30), ECE is computed using 10 equal-width bins from 0 to 1:

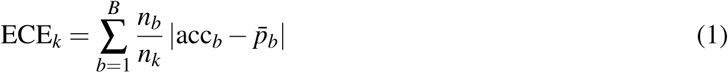

The gap across subgroups is summarized as two values:

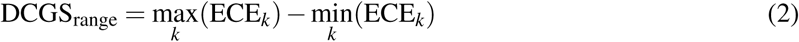

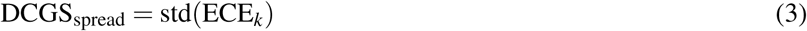

DCGS_range above 0.05 is treated as clinically meaningful. At that threshold, predicted probabilities are at least five percentage points less accurate for the worst-calibrated subgroup than for the best. Bootstrap 95% confidence intervals were estimated from 1,000 stratified resamples. The 0.05 threshold is a proposed convention tied to the clinical decision context: at a biopsy referral cutoff near 25%, a calibration gap of that magnitude shifts a non-trivial fraction of patients across the decision boundary based on demographic group alone.

### Conformal prediction

Nonconformity scores were defined as 1 minus the predicted positive-class probability. Conformal thresholds were fitted from Wisconsin validation nonconformity scores at the 90th quantile (alpha=0.10), targeting 90% marginal coverage. These thresholds were then applied to Wisconsin test and MIMIC evaluation data without adjustment. Demographic Coverage Gap (DCG) was computed as DCG = max*_k_*(cov*_k_*)*−*min*_k_*(cov*_k_*) across racial and gender subgroups with at least 30 members.

### Evaluation metrics

For each model-calibration combination: ECE, maximum calibration error (MCE), Brier Score, Brier Skill Score (where BSS = 1 *−* BS*/*BS_ref_; BS_ref_ is the climatological baseline from outcome prevalence), AUROC, and area under the precision-recall curve (AUPRC), following established prediction model evaluation frameworks [30]. All analyses ran in Python 3.13 with scikit-learn 1.8.0, XGBoost, and SciPy.

## 4 Results and Discussion

### Wisconsin internal calibration

All five models achieved AUROC above 0.98 on the Wisconsin test set (Table 1). XGBoost’s base model had the best ECE at 0.023, followed by RF isotonic at 0.023. Brier Skill Score was positive for every model and calibration condition, confirming each outperformed the naive prevalence baseline. LR base had lower ECE (0.035) than its Platt variant (0.077). That is expected when the base model is already logistic and the calibration sample is small (n=114). SVM Platt produced the highest ECE on Wisconsin at 0.116.

**Table 1:** Wisconsin Diagnostic Breast Cancer internal calibration results (test set, n=114).

| Model | Calibration | ECE | MCE | Brier | BSS | AUROC | AUPRC |
| --- | --- | --- | --- | --- | --- | --- | --- |
| LR | base | 0.035 | 0.431 | 0.024 | 0.896 | 0.9954 | 0.9936 |
| LR | platt | 0.077 | 0.537 | 0.036 | 0.847 | 0.9980 | 0.9966 |
| LR | isotonic | 0.039 | 0.264 | 0.043 | 0.814 | 0.9901 | 0.9725 |
| RF | base | 0.066 | 0.625 | 0.032 | 0.864 | 0.9970 | 0.9951 |
| RF | platt | 0.047 | 0.691 | 0.027 | 0.882 | 0.9959 | 0.9935 |
| RF | isotonic | 0.023 | 0.357 | 0.030 | 0.870 | 0.9917 | 0.9843 |
| SVM | base | 0.070 | 0.586 | 0.039 | 0.833 | 0.9964 | 0.9940 |
| SVM | platt | 0.116 | 0.647 | 0.071 | 0.693 | 0.9964 | 0.9940 |
| SVM | isotonic | 0.040 | 0.651 | 0.041 | 0.826 | 0.9940 | 0.9868 |
| NB | base | 0.076 | 0.792 | 0.074 | 0.683 | 0.9897 | 0.9834 |
| NB | platt | 0.043 | 0.621 | 0.063 | 0.731 | 0.9894 | 0.9828 |
| NB | isotonic | 0.046 | 0.612 | 0.044 | 0.812 | 0.9893 | 0.9817 |
| XGB | base | 0.023 | 0.554 | 0.021 | 0.909 | 0.9934 | 0.9913 |
| XGB | platt | 0.063 | 0.721 | 0.029 | 0.875 | 0.9974 | 0.9957 |
| XGB | isotonic | 0.042 | 0.448 | 0.027 | 0.883 | 0.9982 | 0.9957 |

ECE: Expected Calibration Error. MCE: Maximum Calibration Error. BSS: Brier Skill Score. Calibration: base = no calibration, platt = Platt scaling, isotonic = isotonic regression.

Calibration method and ECE did not move in a consistent direction across models. Isotonic regression reduced ECE for RF and NB while slightly increasing it for LR and XGB. That pattern is expected when calibration and test sets are both small and the base model is already moderately well calibrated. Reliability diagrams for all five models appear in Figures 1-5.

### MIMIC-IV external validation

Calibration degraded sharply on MIMIC (Table 2). AUROC for base, Platt, and isotonic variants fell to 0.45– 0.57 across all five models, confirming that the distributional shift between Wisconsin FNA features and MIMIC triage vitals is severe, as the stress-test design intended. BSS turned negative for most combinations, meaning the calibrated models performed worse on Brier Score than a model predicting a constant 25% prevalence rate. AUPRC for these same combinations clustered near 0.25, which is the random-classifier baseline at 25% positive prevalence. The two metrics agree: base and globally calibrated models carry no useful signal on this cohort.

Subgroup-targeted Platt scaling partially recovered discrimination. AUROC rose to 0.59-0.75 for the subgroup_platt condition, with LR reaching 0.755 and RF reaching 0.745. BSS turned positive for LR (0.051) and RF (0.108), though it stayed negative for SVM and NB. Subgroup-specific recalibration on in-distribution data provides real signal even when the base feature space is mismatched.

ECE on MIMIC varied dramatically. SVM base had an ECE of 0.747. NB base had 0.248. XGB base had 0.181. Global Platt scaling reduced SVM ECE from 0.747 to 0.007 but raised NB ECE from 0.248 to 0.672. Global isotonic produced near-zero ECE for LR and SVM but ECE above 0.22 for XGB. Global calibration methods interact unpredictably with base models on out-of-distribution data. Population reliability curves appear in Figures 6-10.

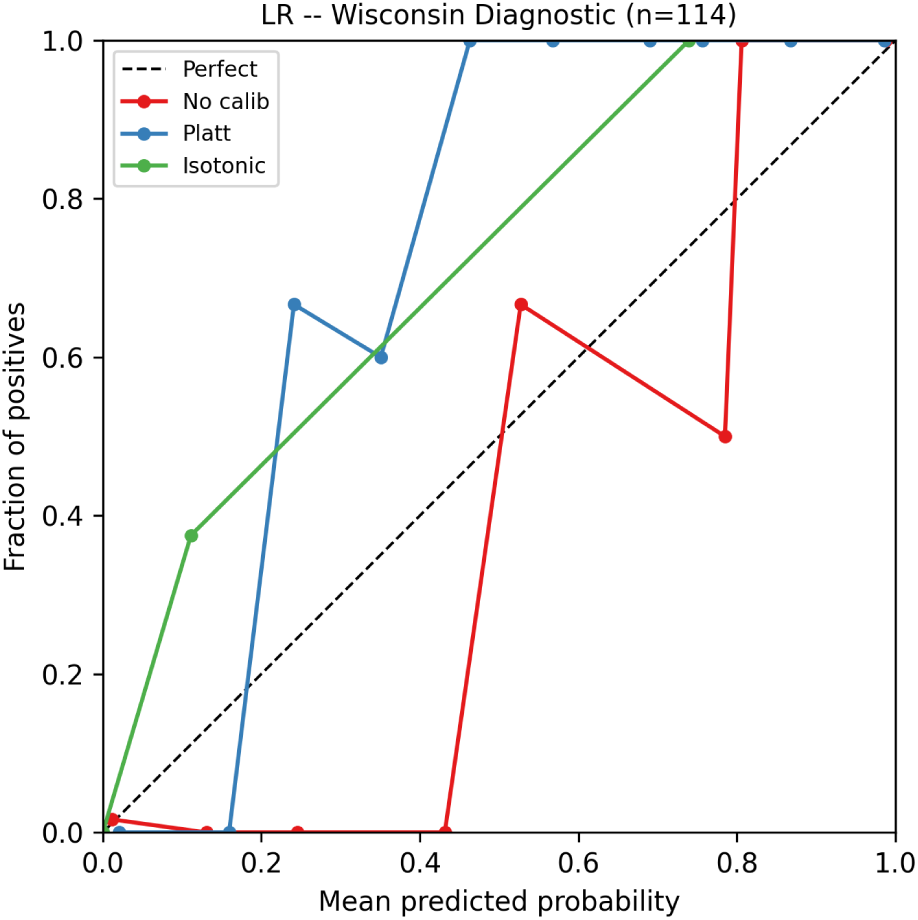
(a) Reliability diagram for LR on the Wisconsin test set (n=114). All three calibration variants are shown. The dashed diagonal represents perfect calibration.

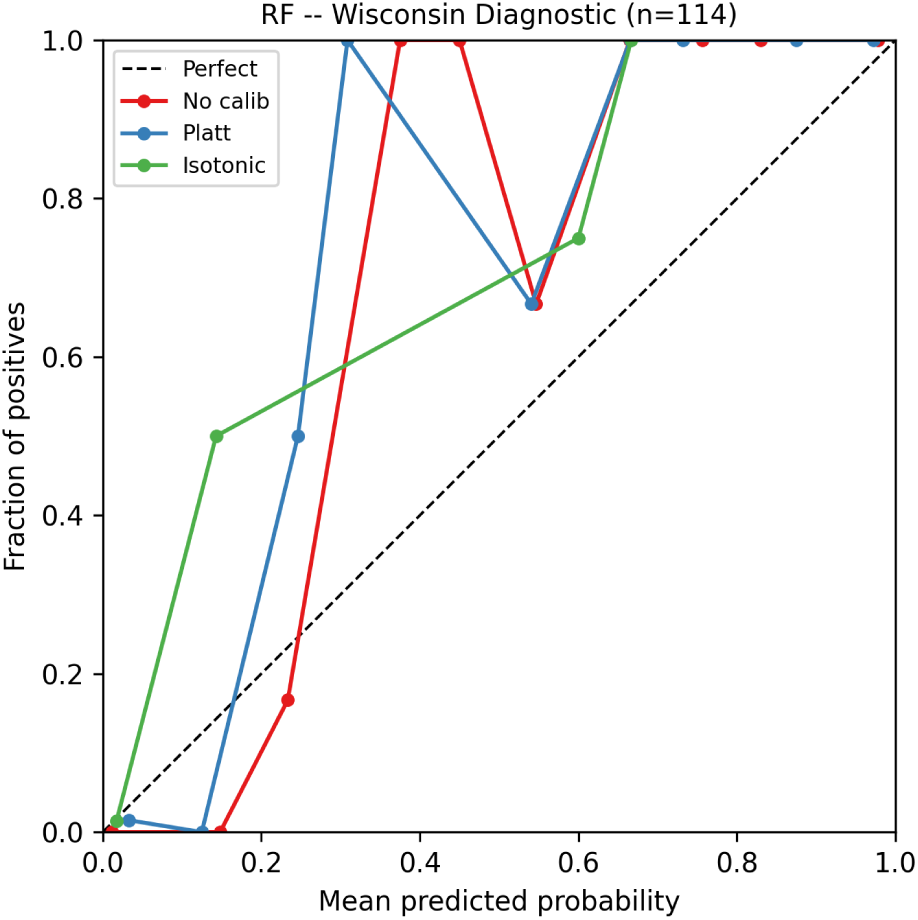
(b) Reliability diagram for RF on the Wisconsin test set (n=114). All three calibration variants are shown. The dashed diagonal represents perfect calibration.

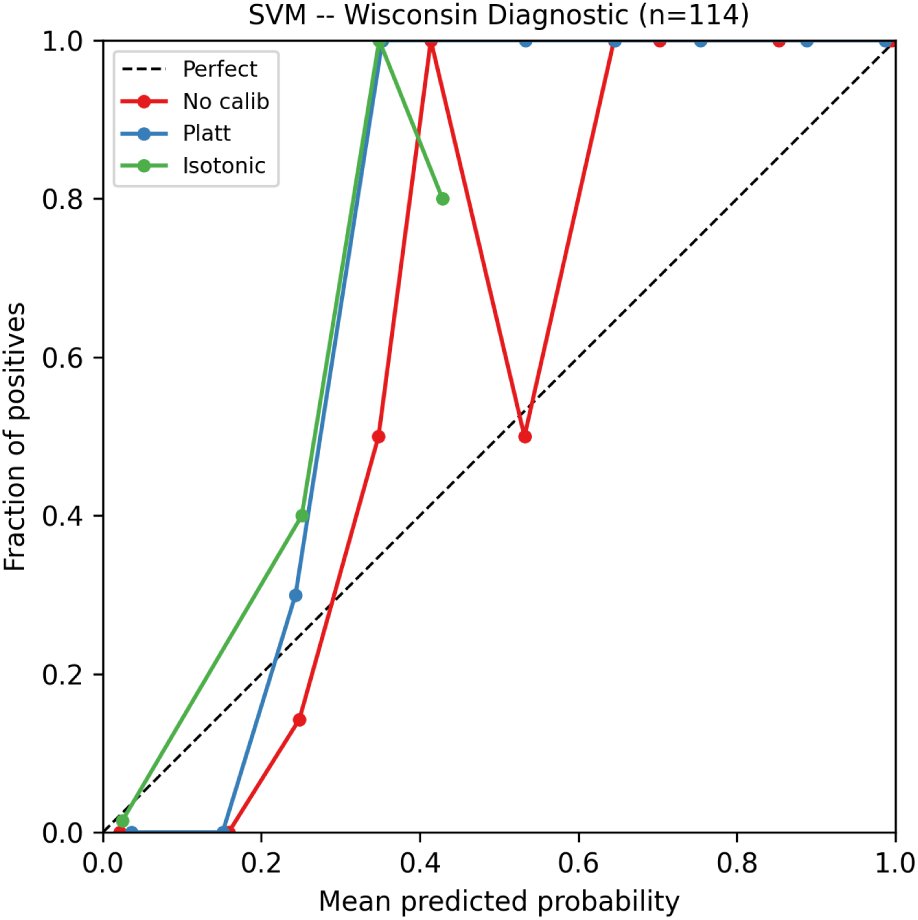
(a) Reliability diagram for SVM on the Wisconsin test set (n=114). All three calibration variants are shown. The dashed diagonal represents perfect calibration.

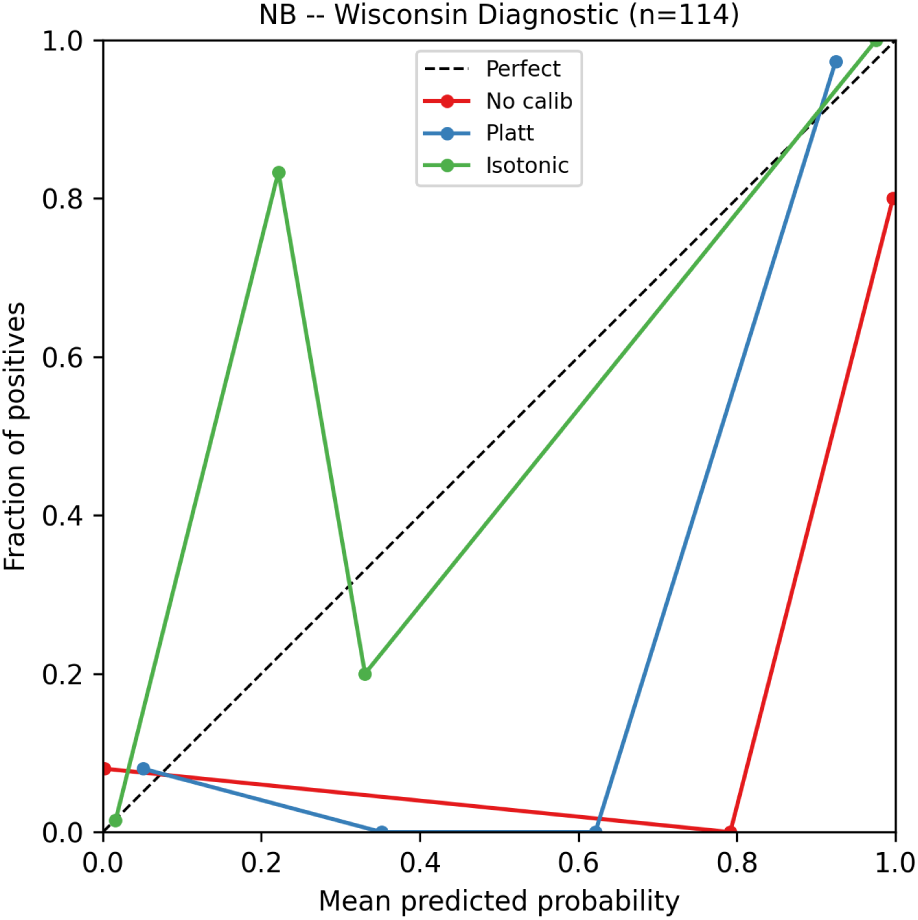
(b) Reliability diagram for NB on the Wisconsin test set (n=114). All three calibration variants are shown. The dashed diagonal represents perfect calibration.

**Table 2:** MIMIC-IV external validation calibration results (evaluation half, n=658).

| Model | Calibration | ECE | Brier | BSS | AUROC | AUPRC |
| --- | --- | --- | --- | --- | --- | --- |
| LR | base | 0.019 | 0.734 | -2.921 | 0.5203 | 0.2571 |
| LR | platt | 0.024 | 0.733 | -2.919 | 0.5223 | 0.2579 |
| LR | isotonic | 0.000 | 0.740 | -2.955 | 0.5071 | 0.2519 |
| LR | subgroup_platt | 0.037 | 0.178 | 0.051 | 0.7545 | 0.3964 |
| RF | base | 0.060 | 0.194 | -0.038 | 0.4553 | 0.2671 |
| RF | platt | 0.076 | 0.194 | -0.035 | 0.4596 | 0.2705 |
| RF | isotonic | 0.060 | 0.188 | -0.006 | 0.5596 | 0.2744 |
| RF | subgroup_platt | 0.072 | 0.167 | 0.108 | 0.7445 | 0.4116 |
| SVM | base | 0.747 | 0.746 | -2.988 | 0.5030 | 0.2504 |
| SVM | platt | 0.007 | 0.737 | -2.940 | 0.5102 | 0.2531 |
| SVM | isotonic | 0.000 | 0.737 | -2.939 | 0.5091 | 0.2527 |
| SVM | subgroup_platt | 0.010 | 0.206 | -0.100 | 0.5936 | 0.3014 |
| NB | base | 0.248 | 0.284 | -0.519 | 0.4757 | 0.2480 |
| NB | platt | 0.672 | 0.640 | -2.421 | 0.5030 | 0.2504 |
| NB | isotonic | 0.722 | 0.710 | -2.796 | 0.5030 | 0.2504 |
| NB | subgroup_platt | 0.047 | 0.201 | -0.072 | 0.6481 | 0.3368 |
| XGB | base | 0.181 | 0.220 | -0.173 | 0.5091 | 0.2527 |
| XGB | platt | 0.208 | 0.230 | -0.232 | 0.5247 | 0.2597 |
| XGB | isotonic | 0.249 | 0.249 | -0.332 | 0.4980 | 0.2492 |
| XGB | subgroup_platt | 0.107 | 0.181 | 0.034 | 0.6690 | 0.3857 |

subgroup\_platt: per-race Platt scaler fitted on MIMIC calibration half (n=658), with global Platt fallback for subgroups with fewer than 30 calibration instances. Negative BSS indicates worse performance than the naive prevalence baseline.

**Figure 3:**
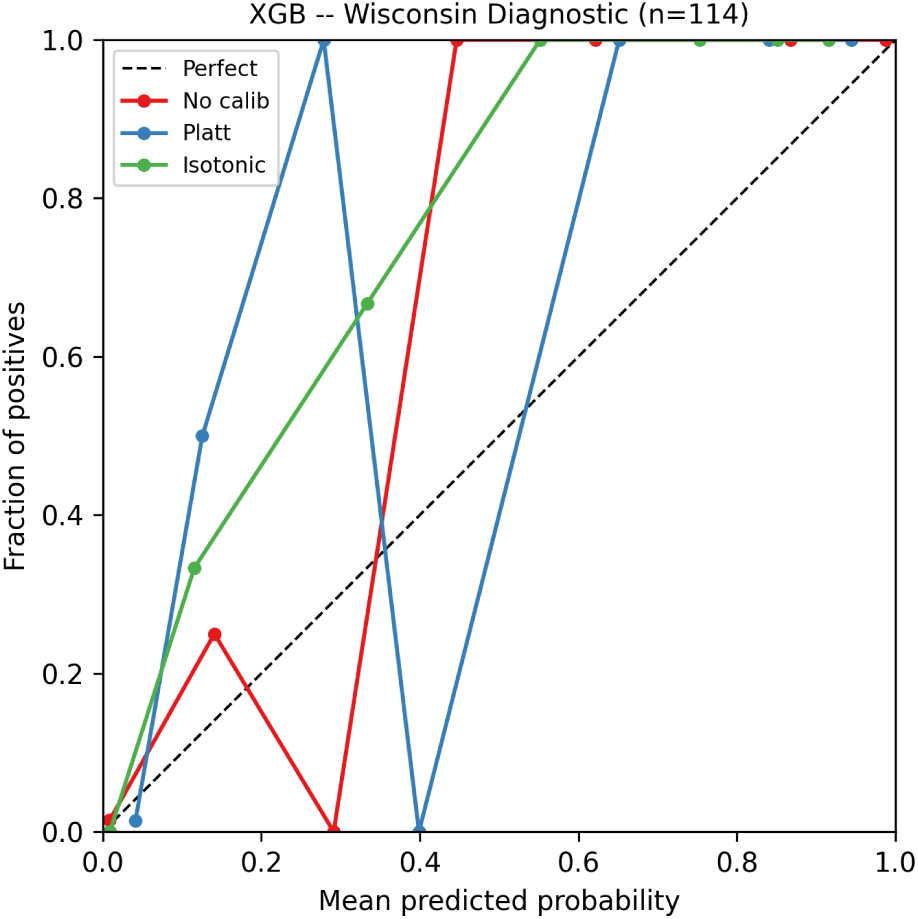
Reliability diagram for XGB on the Wisconsin test set (n=114). All three calibration variants are shown. The dashed diagonal represents perfect calibration.

**Figure 4:**
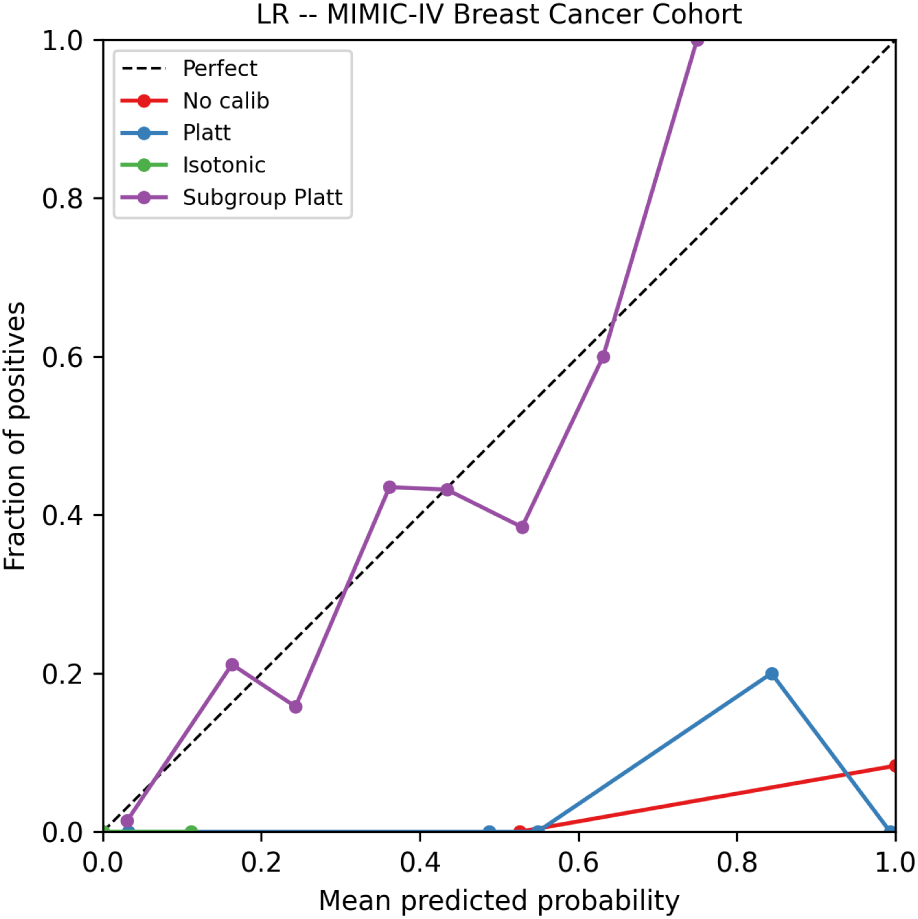
Population-level reliability diagram for LR on the MIMIC-IV evaluation half (n=658). All four calibration variants are shown. The dashed diagonal represents perfect calibration.

### Demographic Calibration Gap Score by race

DCGS exceeded the 0.05 clinical significance threshold in 28 of 40 model-calibration combinations on the race axis (Table 3). Base models showed the largest gaps: NB at 0.195, XGB at 0.191, SVM at 0.176, RF at 0.128, and LR at 0.089. Bootstrap 95% confidence intervals excluded zero for all DCGS values above 0.05.

Global Platt scaling reduced DCGS for SVM (from 0.176 to 0.043) but had negligible effect for XGB (from 0.191 to 0.195). Global isotonic regression produced near-zero DCGS for LR (0.0003) and SVM (0.0009) but left XGB’s race DCGS at 0.195, identical to its uncalibrated baseline. The isotonic calibration that eliminated LR’s race gap did nothing for XGB’s. No globally reliable method emerged from the combinations tested.

The OTHER racial category had the highest ECE in most base model combinations. For NB base, per-group ECE ranged from 0.169 (HISPANIC) to 0.313 (ASIAN). For XGB base, ASIAN had ECE of 0.243 while HISPANIC had 0.101. These patterns shift across models. A single calibration strategy will not equitably correct all subgroups at once. Per-race subgroup reliability diagrams appear in Figures 11-15.

One result demands direct attention. NB with subgroup_platt calibration reaches DCGS_range of 0.602, the highest value in the entire table and more than twice the next-highest subgroup_platt entry. The three other NB calibration conditions hold DCGS between 0.177 and 0.195. Applying subgroup-targeted calibration to NB amplified the racial calibration gap rather than closing it. Two factors interact to produce this. NB’s predicted probabilities are shaped by its naive independence assumption, which generates extreme scores clustered near 0 or 1 on out-of-distribution MIMIC data. The MIMIC calibration half provides roughly 30 patients per racial group at the minimum threshold. Fitting separate Platt scalers to those extreme, sparse distributions pushes subgroup scores in opposite directions. Some groups benefit while others worsen sharply. The NB subgroup_platt result is a practical caution: subgroup-targeted calibration with a poorly behaved base model and small per-group samples amplifies disparity rather than reducing it.

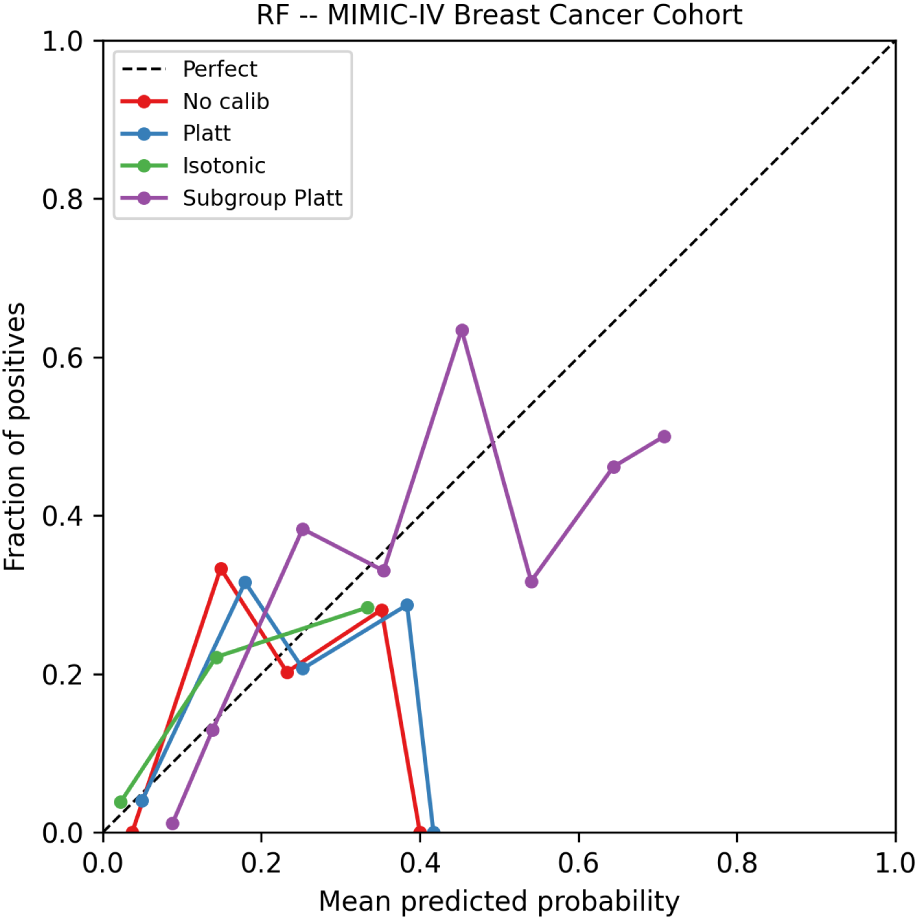
(a) Population-level reliability diagram for RF on the MIMIC-IV evaluation half (n=658). All four calibration variants are shown. The dashed diagonal represents perfect calibration.

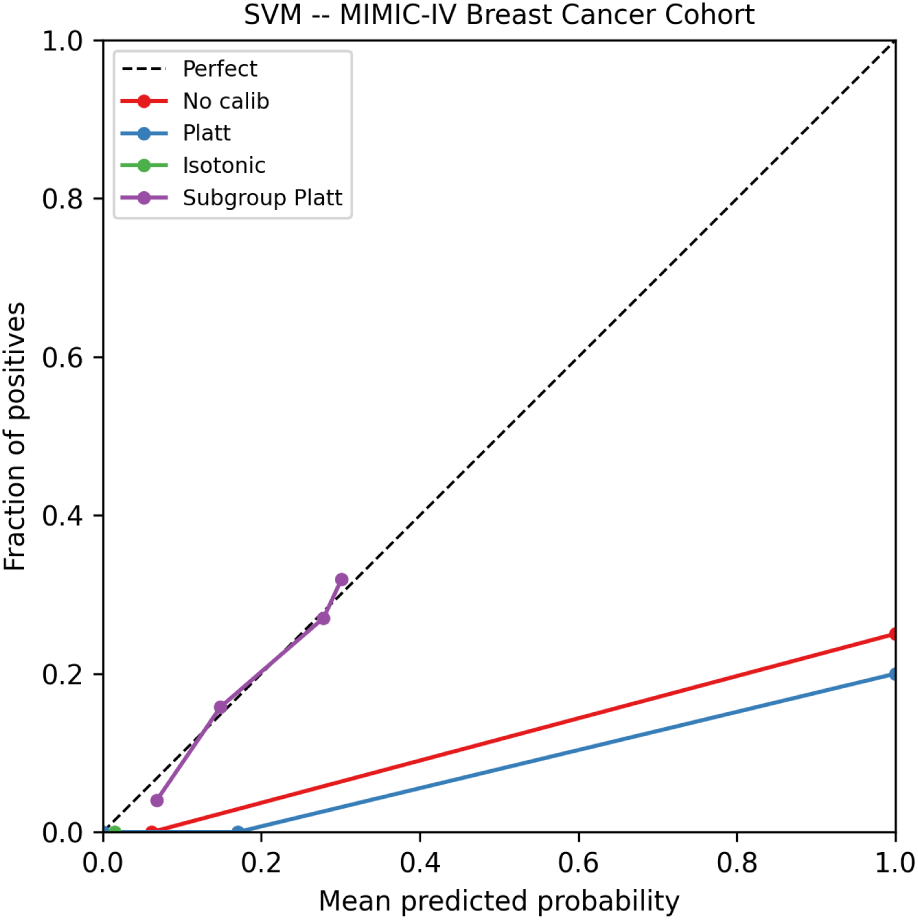
(b) Population-level reliability diagram for SVM on the MIMIC-IV evaluation half (n=658). All four calibration variants are shown. The dashed diagonal represents perfect calibration.

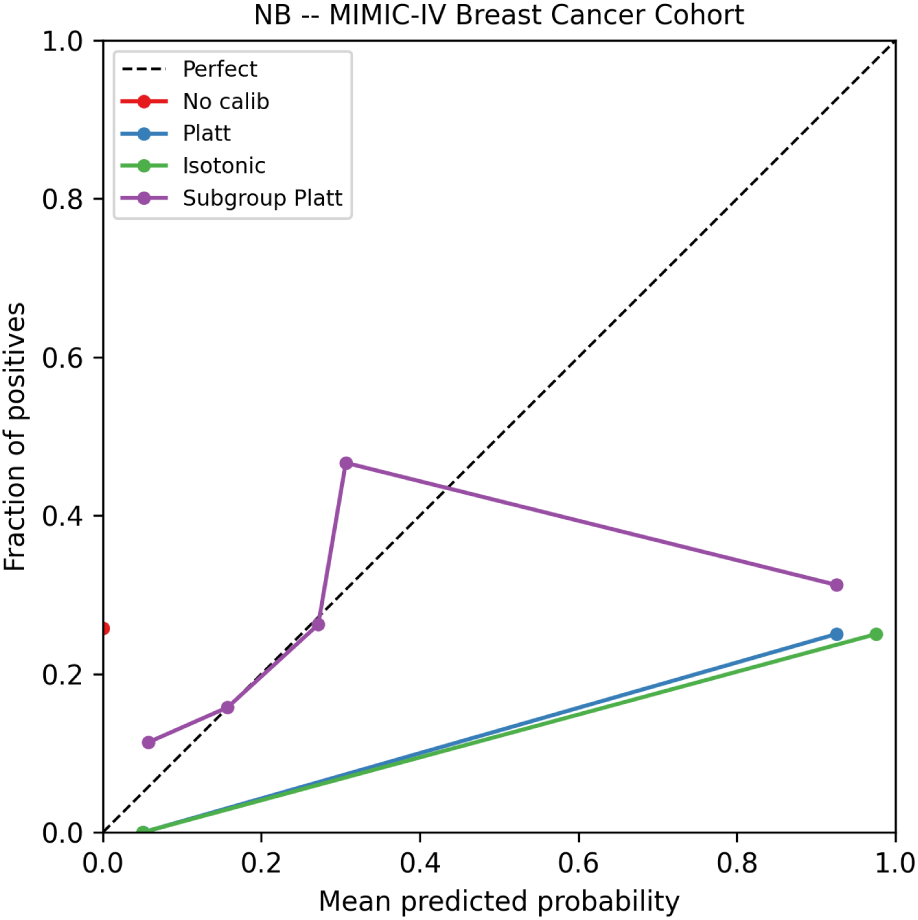
(a) Population-level reliability diagram for NB on the MIMIC-IV evaluation half (n=658). All four calibration variants are shown. The dashed diagonal represents perfect calibration.

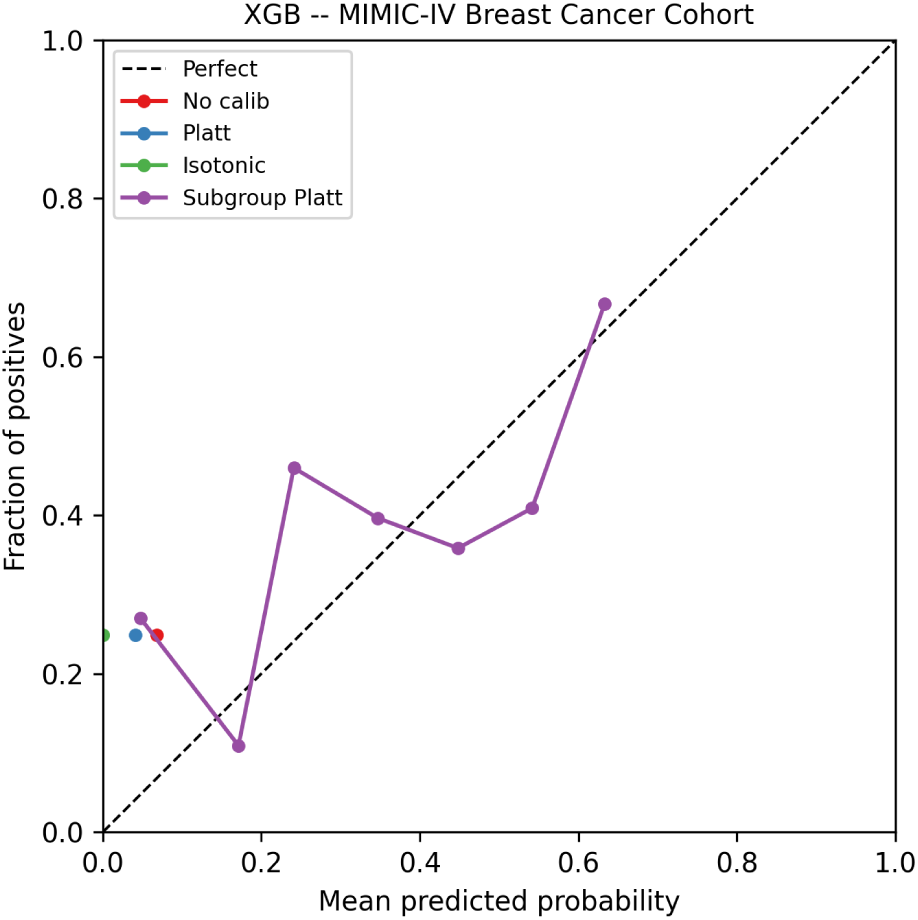
(b) Population-level reliability diagram for XGB on the MIMIC-IV evaluation half (n=658). All four calibration variants are shown. The dashed diagonal represents perfect calibration.

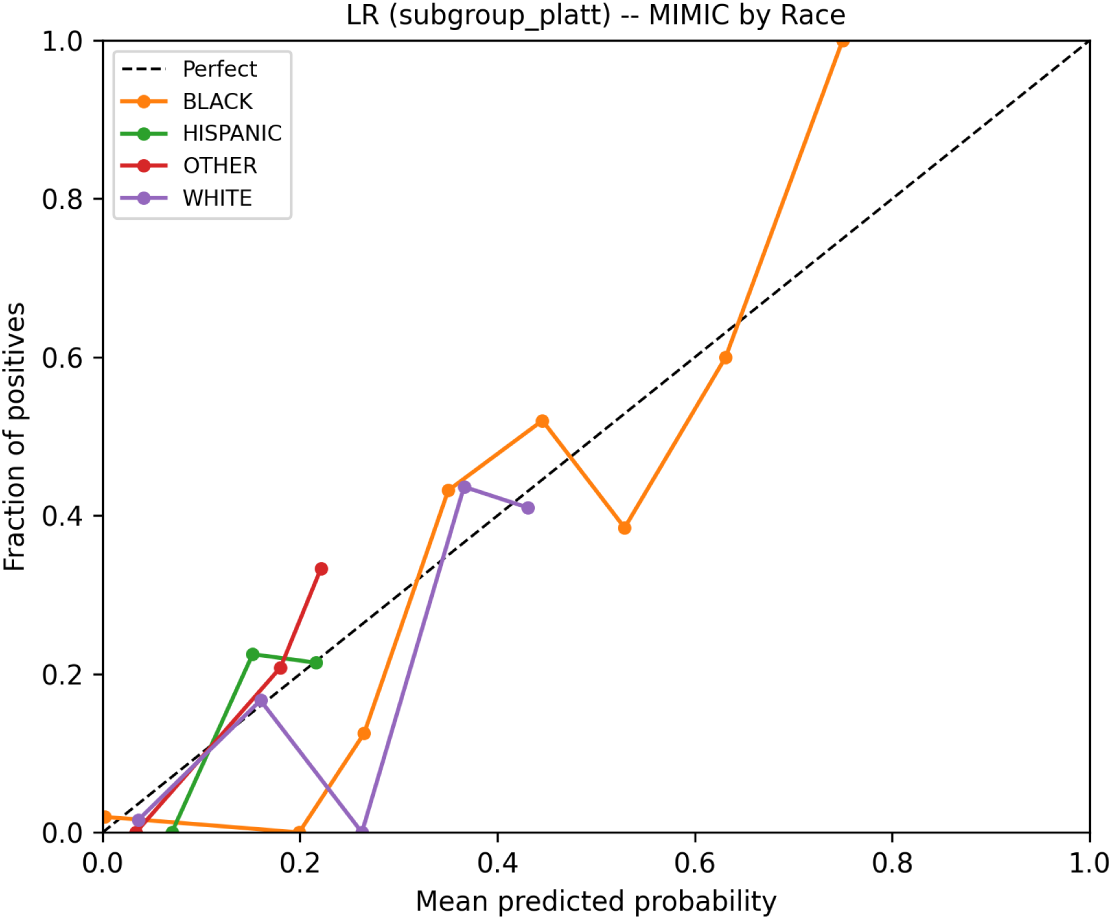
(a) Subgroup reliability diagram by race for LR (subgroup_platt variant) on the MIMIC-IV evaluation half. Each line represents one racial subgroup with at least 50 patients. The dashed diagonal represents perfect calibration.

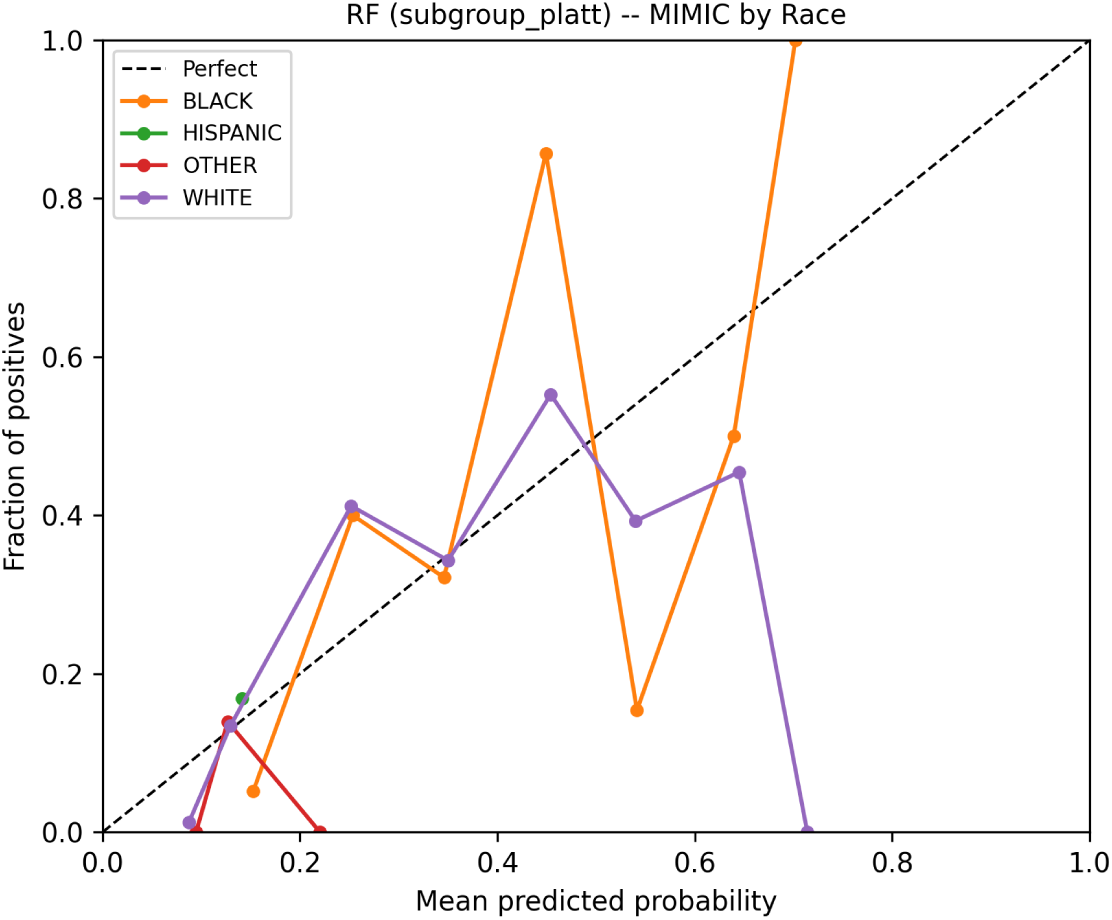
(b) Subgroup reliability diagram by race for RF (subgroup_platt variant) on the MIMIC-IV evaluation half. Each line represents one racial subgroup with at least 50 patients. The dashed diagonal represents perfect calibration.

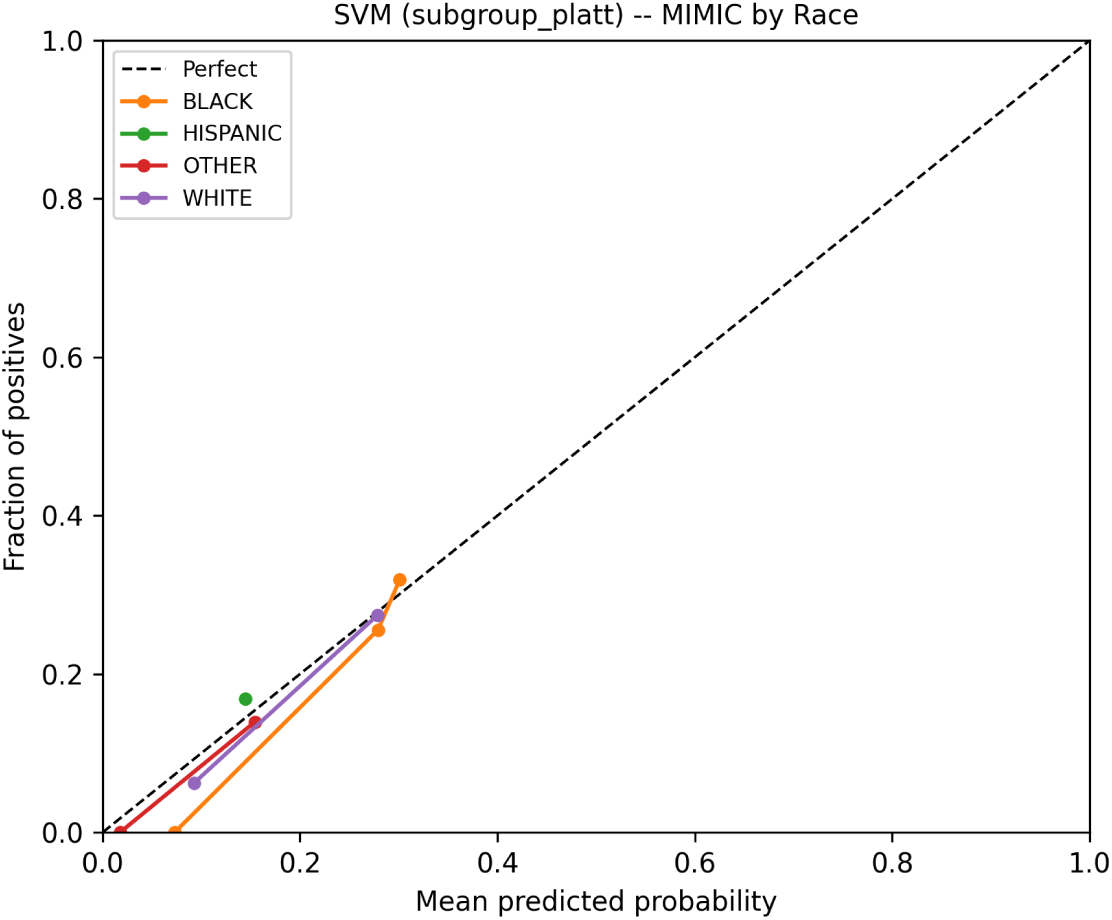
(a) Subgroup reliability diagram by race for SVM (subgroup_platt variant) on the MIMIC-IV evaluation half. Each line represents one racial subgroup with at least 50 patients. The dashed diagonal represents perfect calibration.

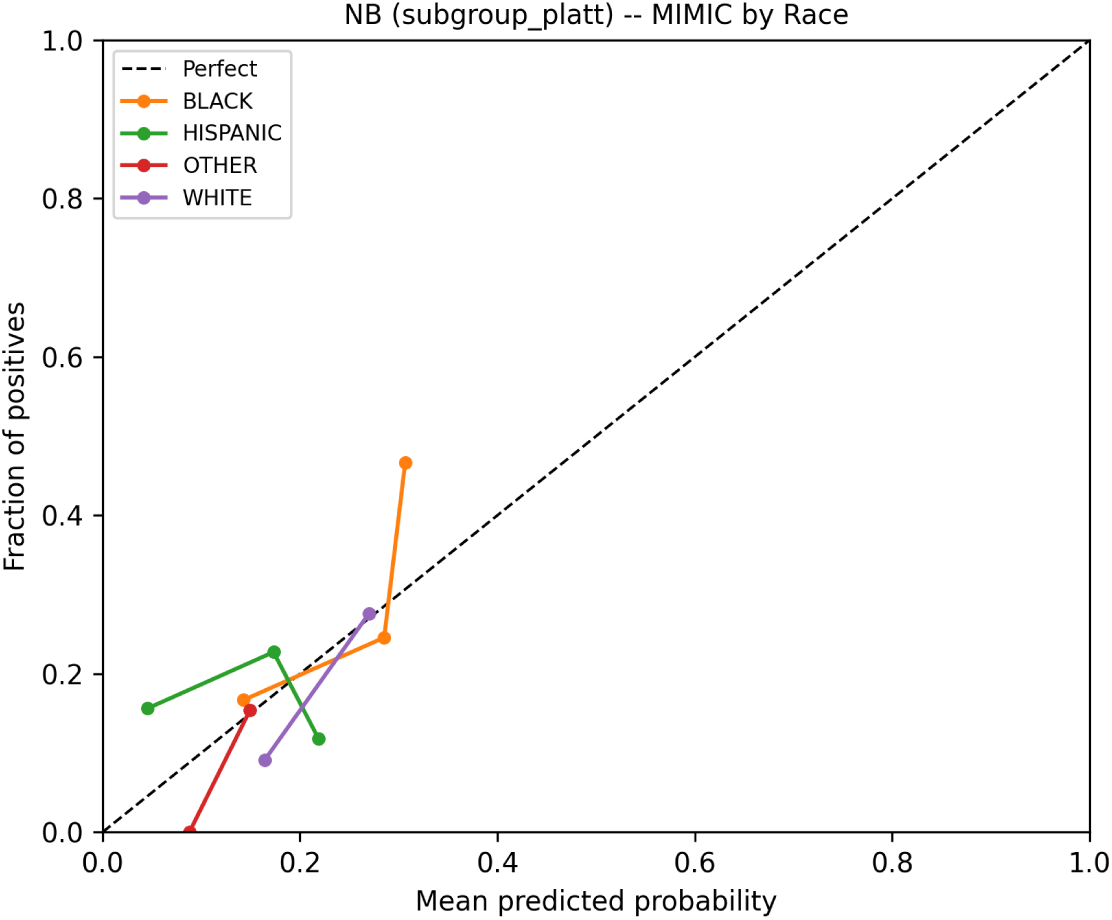
(b) Subgroup reliability diagram by race for NB (subgroup_platt variant) on the MIMIC-IV evaluation half. Each line represents one racial subgroup with at least 50 patients. The dashed diagonal represents perfect calibration.

**Table 3:**
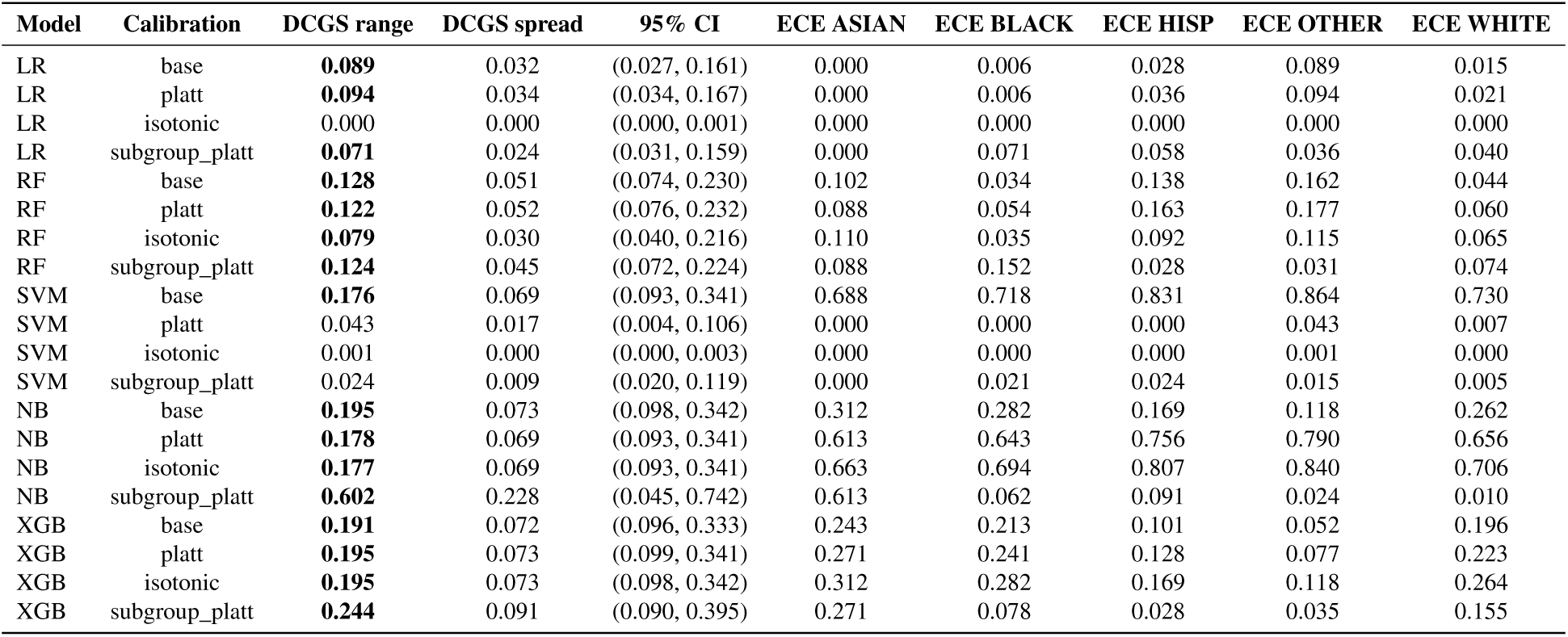
DCGS by race across model and calibration conditions (MIMIC-IV evaluation half, n=658). Values exceeding the 0.05 clinical significance threshold are bolded.

| Model | Calibration | DCGS range | DCGS spread | 95% CI | ECE ASIAN | ECE BLACK | ECE HISP | ECE OTHER | ECE WHITE |
| --- | --- | --- | --- | --- | --- | --- | --- | --- | --- |
| LR | base | **0.089** | 0.032 | (0.027, 0.161) | 0.000 | 0.006 | 0.028 | 0.089 | 0.015 |
| LR | platt | **0.094** | 0.034 | (0.034, 0.167) | 0.000 | 0.006 | 0.036 | 0.094 | 0.021 |
| LR | isotonic | 0.000 | 0.000 | (0.000, 0.001) | 0.000 | 0.000 | 0.000 | 0.000 | 0.000 |
| LR | subgroup_platt | **0.071** | 0.024 | (0.031, 0.159) | 0.000 | 0.071 | 0.058 | 0.036 | 0.040 |
| RF | base | **0.128** | 0.051 | (0.074, 0.230) | 0.102 | 0.034 | 0.138 | 0.162 | 0.044 |
| RF | platt | **0.122** | 0.052 | (0.076, 0.232) | 0.088 | 0.054 | 0.163 | 0.177 | 0.060 |
| RF | isotonic | **0.079** | 0.030 | (0.040, 0.216) | 0.110 | 0.035 | 0.092 | 0.115 | 0.065 |
| RF | subgroup_platt | **0.124** | 0.045 | (0.072, 0.224) | 0.088 | 0.152 | 0.028 | 0.031 | 0.074 |
| SVM | base | **0.176** | 0.069 | (0.093, 0.341) | 0.688 | 0.718 | 0.831 | 0.864 | 0.730 |
| SVM | platt | 0.043 | 0.017 | (0.004, 0.106) | 0.000 | 0.000 | 0.000 | 0.043 | 0.007 |
| SVM | isotonic | 0.001 | 0.000 | (0.000, 0.003) | 0.000 | 0.000 | 0.000 | 0.001 | 0.000 |
| SVM | subgroup_platt | 0.024 | 0.009 | (0.020, 0.119) | 0.000 | 0.021 | 0.024 | 0.015 | 0.005 |
| NB | base | **0.195** | 0.073 | (0.098, 0.342) | 0.312 | 0.282 | 0.169 | 0.118 | 0.262 |
| NB | platt | **0.178** | 0.069 | (0.093, 0.341) | 0.613 | 0.643 | 0.756 | 0.790 | 0.656 |
| NB | isotonic | **0.177** | 0.069 | (0.093, 0.341) | 0.663 | 0.694 | 0.807 | 0.840 | 0.706 |
| NB | subgroup_platt | **0.602** | 0.228 | (0.045, 0.742) | 0.613 | 0.062 | 0.091 | 0.024 | 0.010 |
| XGB | base | **0.191** | 0.072 | (0.096, 0.333) | 0.243 | 0.213 | 0.101 | 0.052 | 0.196 |
| XGB | platt | **0.195** | 0.073 | (0.099, 0.341) | 0.271 | 0.241 | 0.128 | 0.077 | 0.223 |
| XGB | isotonic | **0.195** | 0.073 | (0.098, 0.342) | 0.312 | 0.282 | 0.169 | 0.118 | 0.264 |
| XGB | subgroup_platt | **0.244** | 0.091 | (0.090, 0.395) | 0.271 | 0.078 | 0.028 | 0.035 | 0.155 |

### Demographic Calibration Gap Score by gender

Gender DCGS followed a different pattern than race (Table 3b). LR held gender DCGS below 0.05 across all four calibration conditions, suggesting its probability outputs are reasonably consistent across female and male patients on this cohort. RF and SVM base models had large gender DCGS: 0.192 and 0.379 respectively. SVM Platt scaling brought gender DCGS from 0.379 to 0.011. SVM isotonic reduced it further to 0.0003. RF isotonic calibration reduced gender DCGS to 0.085, still above the threshold.

SVM Platt scaling fixed the gender gap but left race DCGS at 0.043. The same calibration method addressed one axis of equity while barely moving the other. Single-axis reporting misses exactly this kind of partial correction. Per-gender subgroup reliability diagrams appear in Figures 16-20.

The SVM base gender gap at 0.379 warrants explanation. ECE for male patients reached 0.988, near the theoretical maximum. SVM probability estimates on out-of-distribution data saturate the sigmoid transformation, pushing scores toward 0 or 1. The MIMIC breast cancer cohort is predominantly female. Male patients occupy a minority sub-distribution where SVM confidence scores are almost universally wrong. The resulting male ECE of 0.988 against a female ECE of 0.608 produces the large gender DCGS. Platt scaling on the full population corrected this substantially, bringing gender DCGS from 0.379 to 0.011.

### Conformal prediction coverage

Conformal thresholds fitted on Wisconsin validation data transferred poorly to MIMIC (Table 4). Wisconsin test coverage ranged from 0.37 to 0.53, below the 90% target for all models. That shortfall traces to the coverage metric’s construction. Prediction sets include the positive class when the nonconformity score falls below the fitted threshold. A negative test sample, where the true label is zero, has no pathway to coverage under this single-class scheme. Wisconsin test coverage therefore approximates the positive class fraction in the test partition, roughly 37%. Models with thresholds below 1.0 extend coverage toward 0.53 by including the positive-class prediction for a wider range of samples. On MIMIC, coverage clustered near 0.249 for most model-variant combinations. The collapse to roughly 25% corresponds to the label prevalence in the MIMIC evaluation half: the conformal threshold was set so high that prediction sets nearly always excluded the positive class. The two failures have different causes. The Wisconsin shortfall is structural to the single-class implementation; the MIMIC collapse is driven by distributional shift.

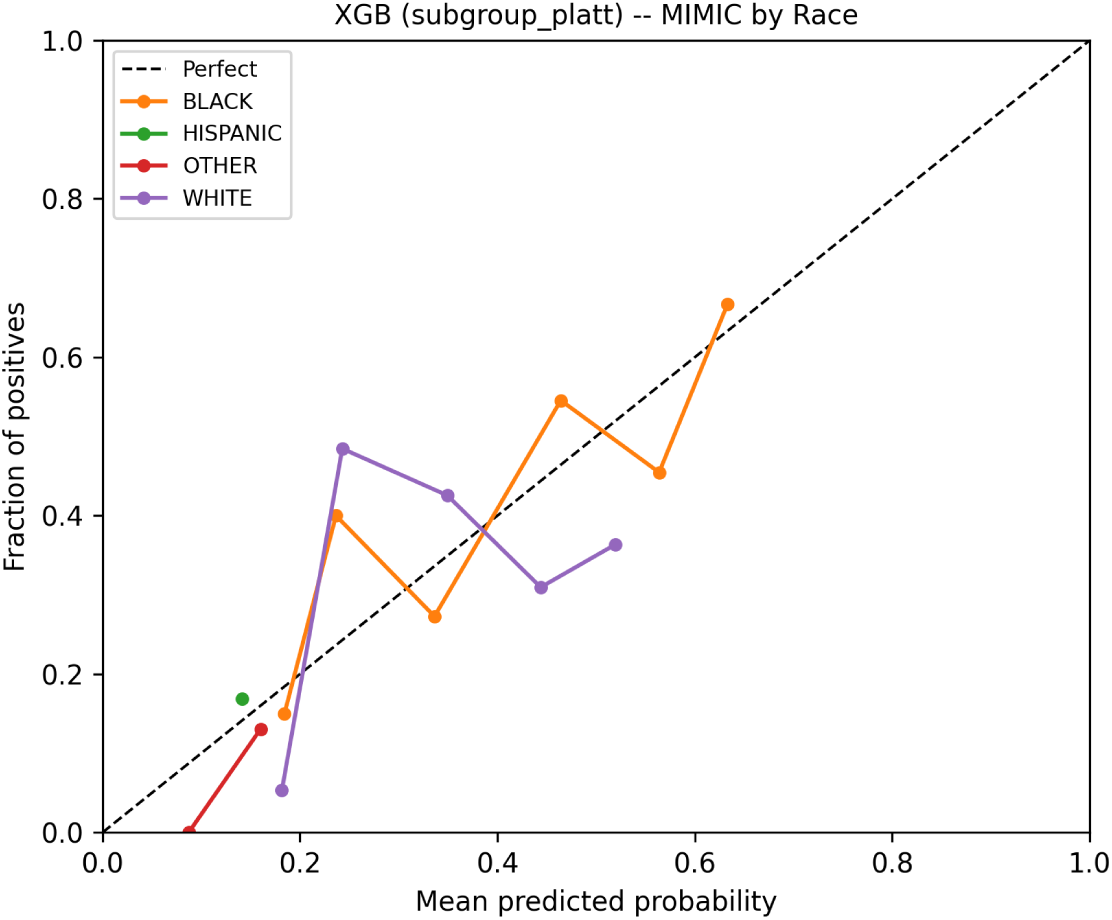
(a) Subgroup reliability diagram by race for XGB (subgroup_platt variant) on the MIMIC-IV evaluation half. Each line represents one racial subgroup with at least 50 patients. The dashed diagonal represents perfect calibration.

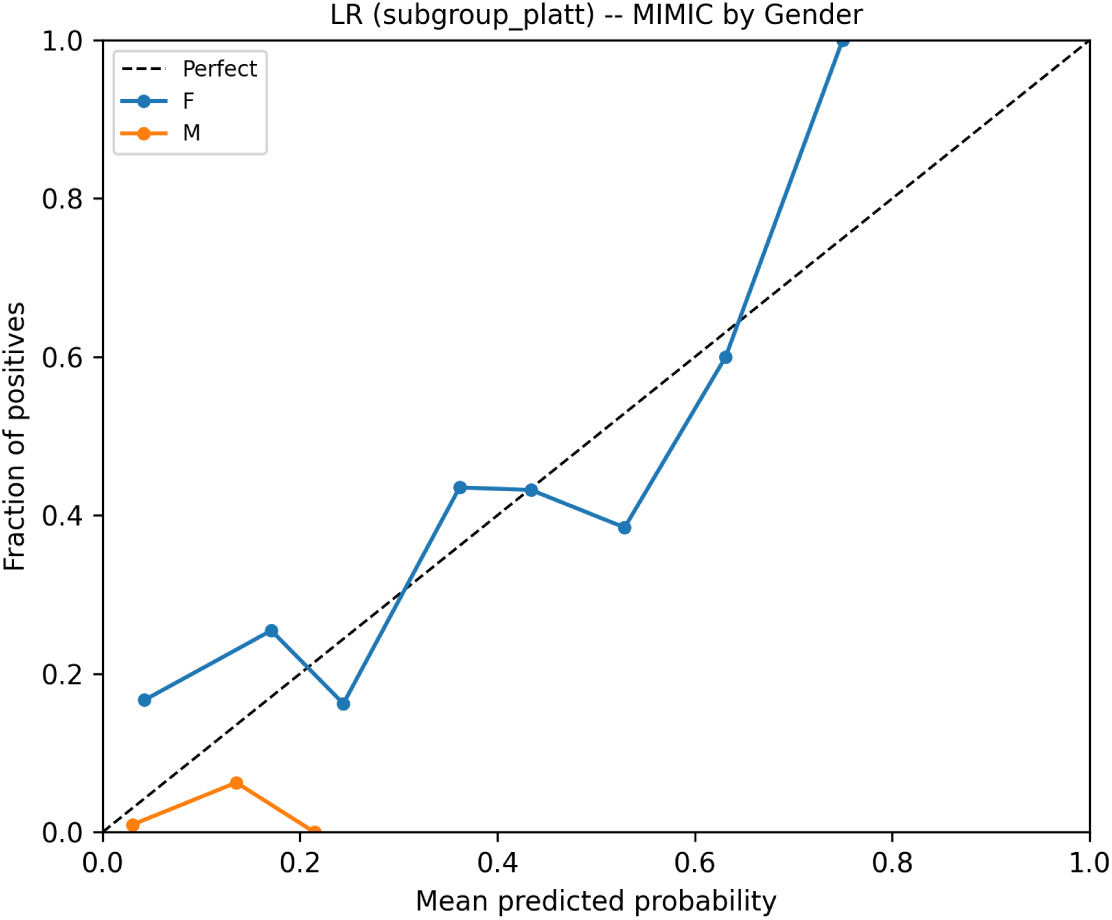
(b) Subgroup reliability diagram by gender for LR (subgroup_platt variant) on the MIMIC-IV evaluation half. Lines represent Female (F) and Male (M) subgroups. The dashed diagonal represents perfect calibration.

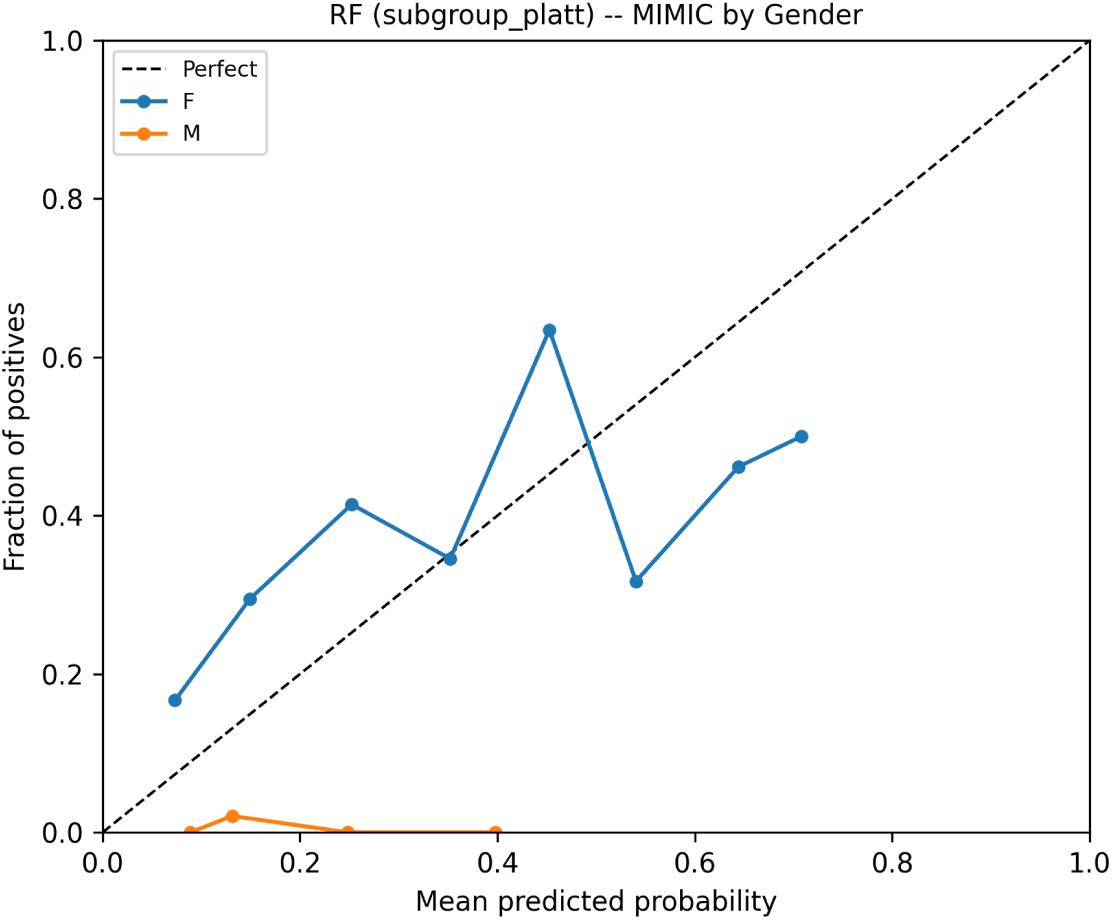
(a) Subgroup reliability diagram by gender for RF (subgroup_platt variant) on the MIMIC-IV evaluation half. Lines represent Female (F) and Male (M) subgroups. The dashed diagonal represents perfect calibration.

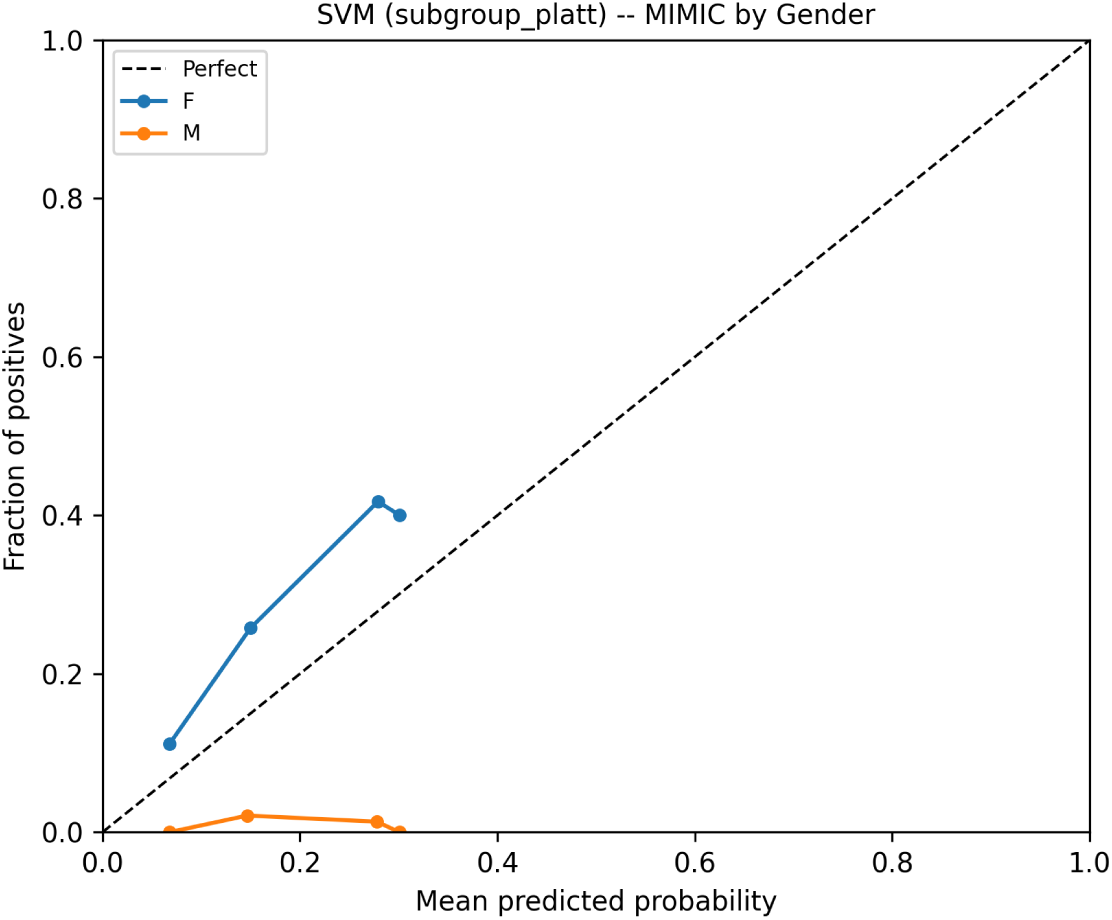
(b) Subgroup reliability diagram by gender for SVM (subgroup_platt variant) on the MIMIC-IV evaluation half. Lines represent Female (F) and Male (M) subgroups. The dashed diagonal represents perfect calibration.

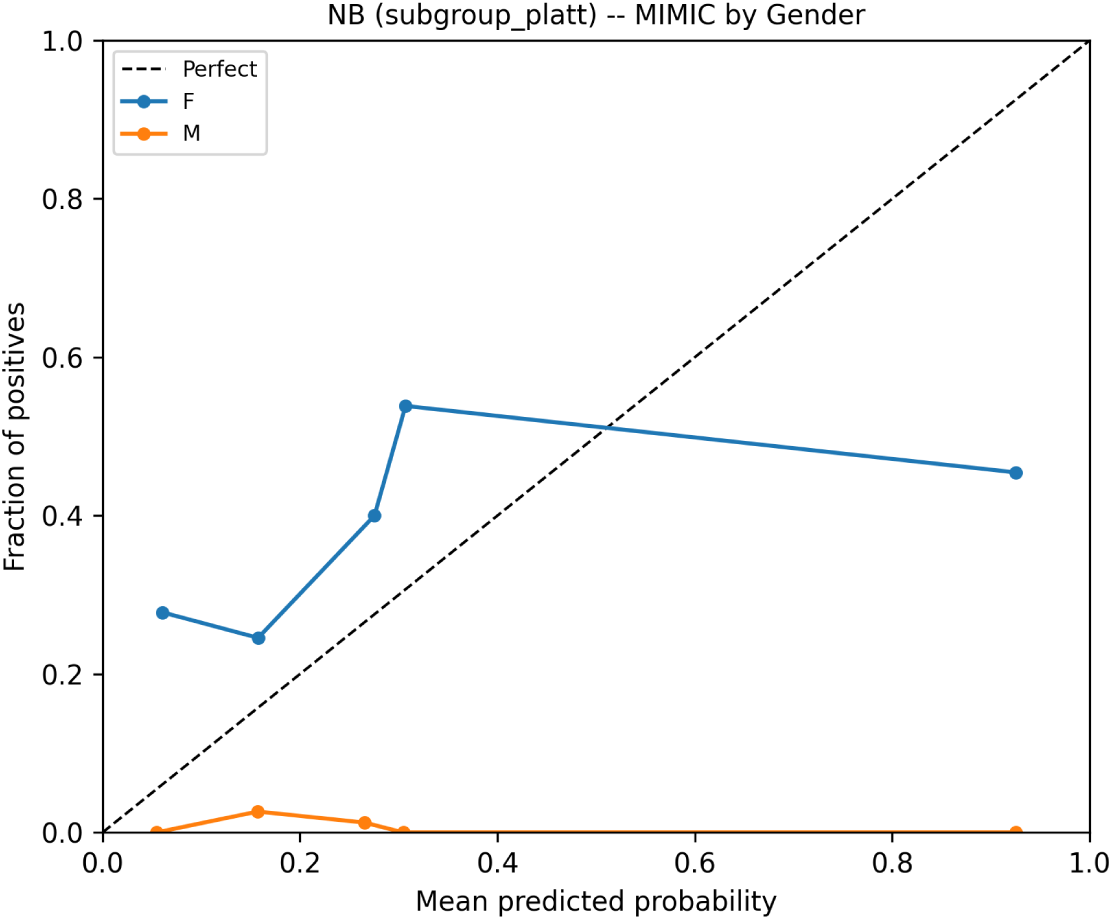
(a) Subgroup reliability diagram by gender for NB (subgroup_platt variant) on the MIMIC-IV evaluation half. Lines represent Female (F) and Male (M) subgroups. The dashed diagonal represents perfect calibration.

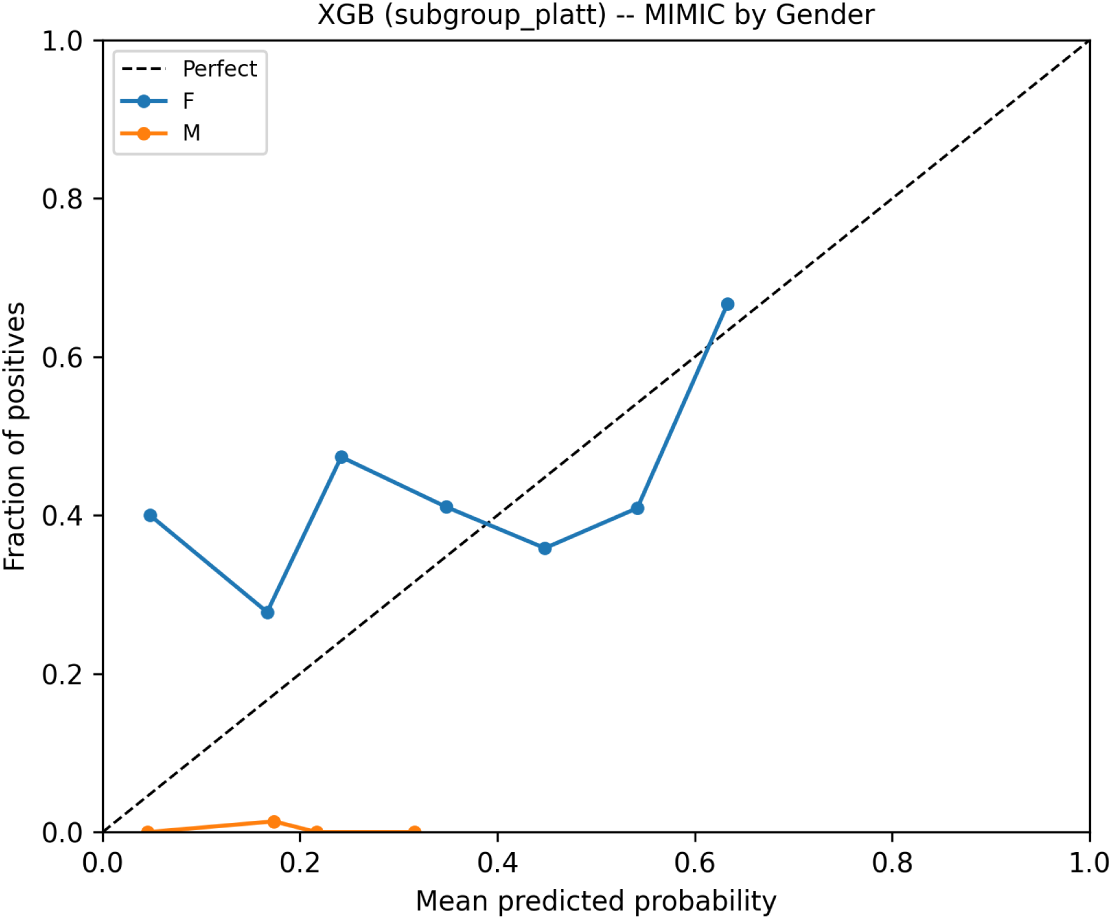
(b) Subgroup reliability diagram by gender for XGB (subgroup_platt variant) on the MIMIC-IV evaluation half. Lines represent Female (F) and Male (M) subgroups. The dashed diagonal represents perfect calibration.

**Table 4:** DCGS by gender across model and calibration conditions (MIMIC-IV evaluation half, n=658). Values exceeding the 0.05 clinical significance threshold are bolded.

| Model | Calibration | DCGS range | DCGS spread | 95% CI | ECE Female | ECE Male |
| --- | --- | --- | --- | --- | --- | --- |
| LR | base | 0.016 | 0.008 | (0.001, 0.042) | 0.013 | 0.029 |
| LR | platt | 0.036 | 0.018 | (0.010, 0.064) | 0.011 | 0.047 |
| LR | isotonic | 0.001 | 0.000 | (0.000, 0.001) | 0.000 | 0.001 |
| LR | subgroup_platt | 0.029 | 0.015 | (0.006, 0.086) | 0.053 | 0.024 |
| RF | base | **0.192** | 0.096 | (0.138, 0.240) | 0.099 | 0.291 |
| RF | platt | **0.236** | 0.118 | (0.181, 0.274) | 0.064 | 0.300 |
| RF | isotonic | **0.085** | 0.043 | (0.031, 0.135) | 0.130 | 0.215 |
| RF | subgroup_platt | 0.012 | 0.006 | (0.002, 0.069) | 0.126 | 0.114 |
| SVM | base | **0.379** | 0.190 | (0.330, 0.428) | 0.608 | 0.988 |
| SVM | platt | 0.011 | 0.005 | (0.003, 0.022) | 0.011 | 0.000 |
| SVM | isotonic | 0.000 | 0.000 | (0.000, 0.001) | 0.000 | 0.000 |
| SVM | subgroup_platt | **0.096** | 0.048 | (0.045, 0.141) | 0.119 | 0.215 |
| NB | base | **0.370** | 0.185 | (0.320, 0.421) | 0.383 | 0.013 |
| NB | platt | **0.379** | 0.190 | (0.329, 0.428) | 0.534 | 0.913 |
| NB | isotonic | **0.379** | 0.190 | (0.330, 0.428) | 0.584 | 0.963 |
| NB | subgroup_platt | **0.093** | 0.046 | (0.037, 0.143) | 0.148 | 0.241 |
| XGB | base | **0.262** | 0.131 | (0.212, 0.313) | 0.317 | 0.055 |
| XGB | platt | **0.316** | 0.158 | (0.266, 0.368) | 0.344 | 0.028 |
| XGB | isotonic | **0.373** | 0.186 | (0.322, 0.424) | 0.385 | 0.012 |
| XGB | subgroup_platt | 0.001 | 0.001 | (0.001, 0.055) | 0.155 | 0.157 |

Racial DCG on MIMIC exceeded 0.15 for all 20 model-variant combinations. The largest racial DCG was 0.41 for LR subgroup_platt. Gender DCG ranged from 0.14 to 0.43. Prediction set guarantees derived from a homogeneous training population do not transfer equitably to diverse external cohorts. DCGS heatmaps for race and gender appear in Figures 21-24.

### Why global calibration fails for equity

A population-level ECE below 0.05 coexists with a DCGS above 0.19. These experiments confirmed the point directly. Across five model types and three standard calibration methods, the racial calibration gap on the MIMIC cohort exceeded 0.05 in 28 of 40 combinations. Platt and isotonic calibration did not reliably bring DCGS below that threshold.

The mechanism is straightforward. A global calibration function shifts predicted probabilities across the full population to reduce aggregate ECE. When one racial group receives systematically higher base model scores than another, the global function adjusts for the population-weighted average. The result undercorrects for one group and overcorrects for another. The subgroup gaps survive.

One apparent exception is worth examining. LR with isotonic calibration achieved DCGS_race of 0.0003. That is not a calibration success in any real sense. Isotonic regression, fitted on 114 Wisconsin validation patients and applied to completely different MIMIC triage features, produces highly concentrated probability outputs. All subgroups end up with near-identical predicted values, so per-group ECEs appear similar. They are equally wrong, not equally right.

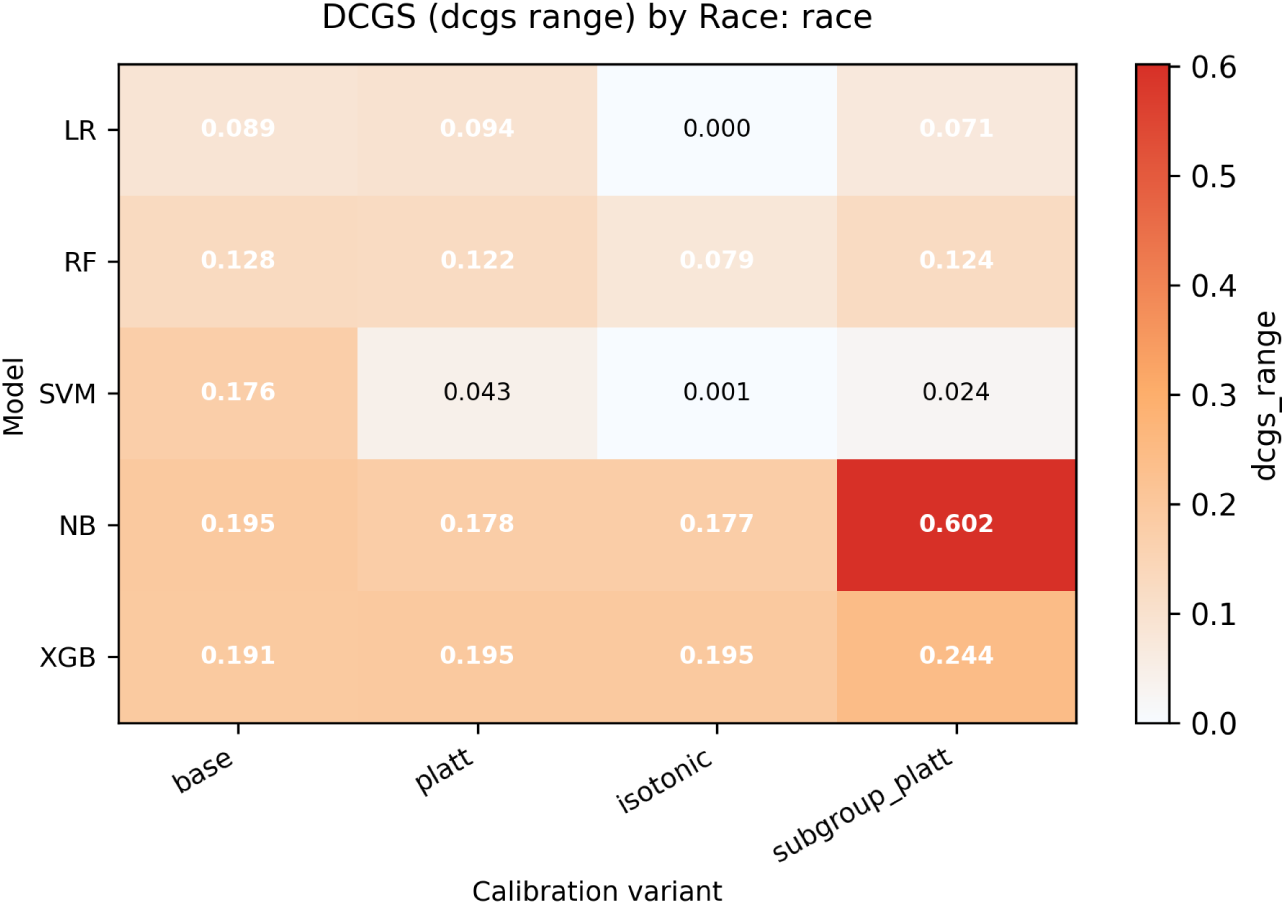
(a) DCGS range heatmap by race (MIMIC-IV evaluation half). Rows are model types. Columns are calibration variants. Cell values show DCGS_range across racial subgroups. Darker shading and bold text indicate values above the 0.05 clinical significance threshold.

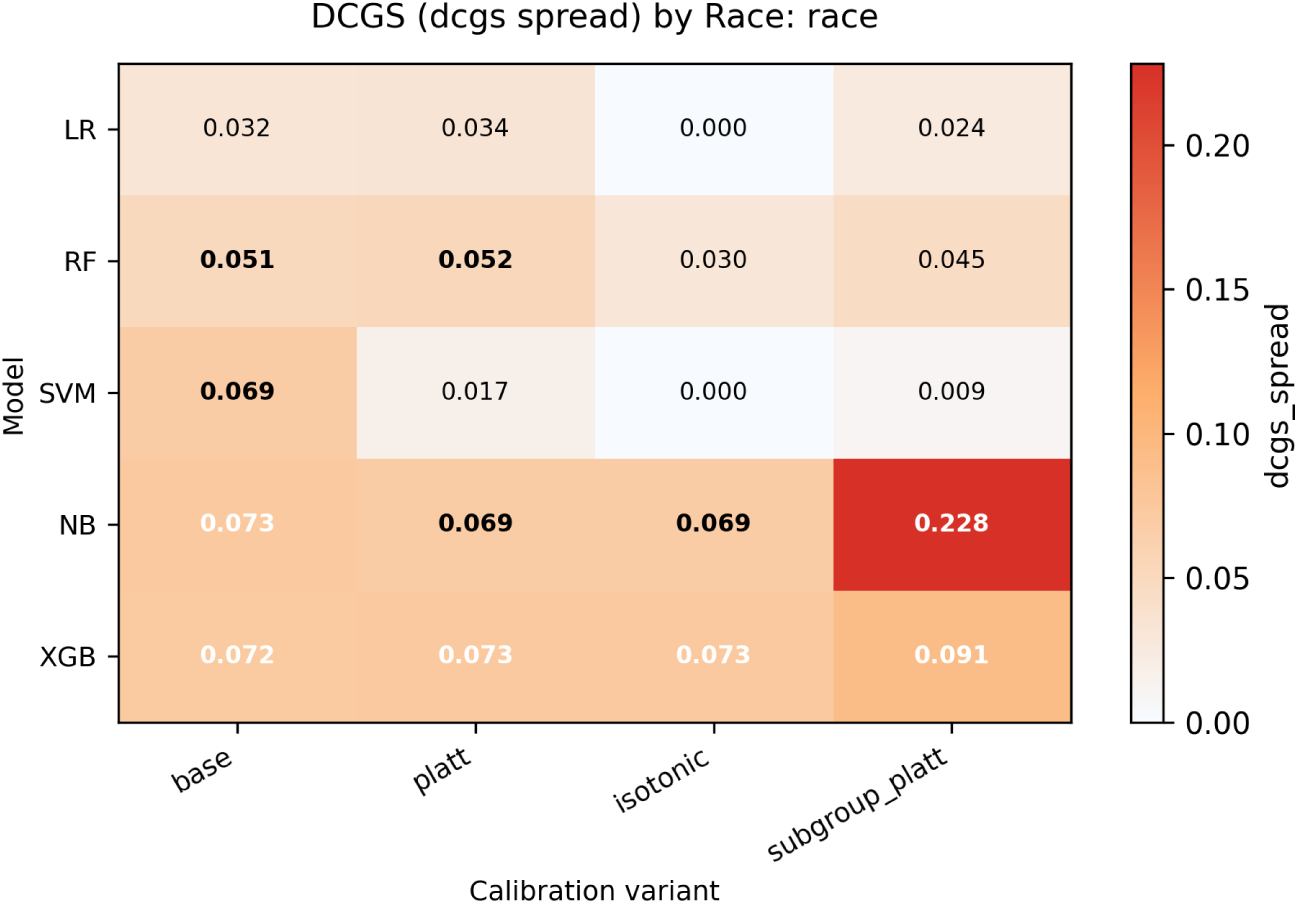
(b) DCGS spread heatmap by race. Cell values show the standard deviation of ECE across racial subgroups, capturing intermediate calibration gaps not visible in the range metric alone.

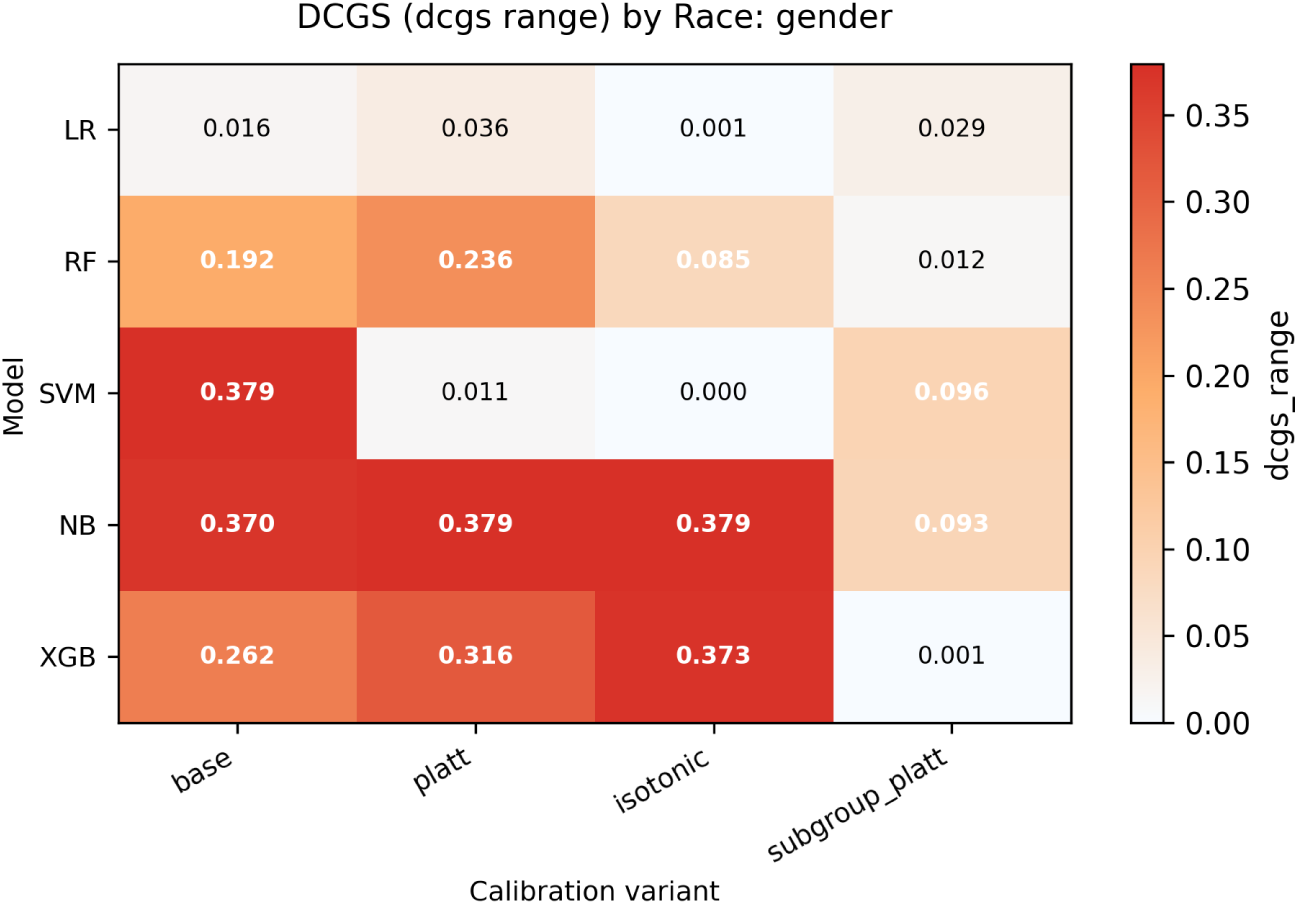
(a) DCGS range heatmap by gender (MIMIC-IV evaluation half). Cell values show DCGS_range across Female and Male subgroups.

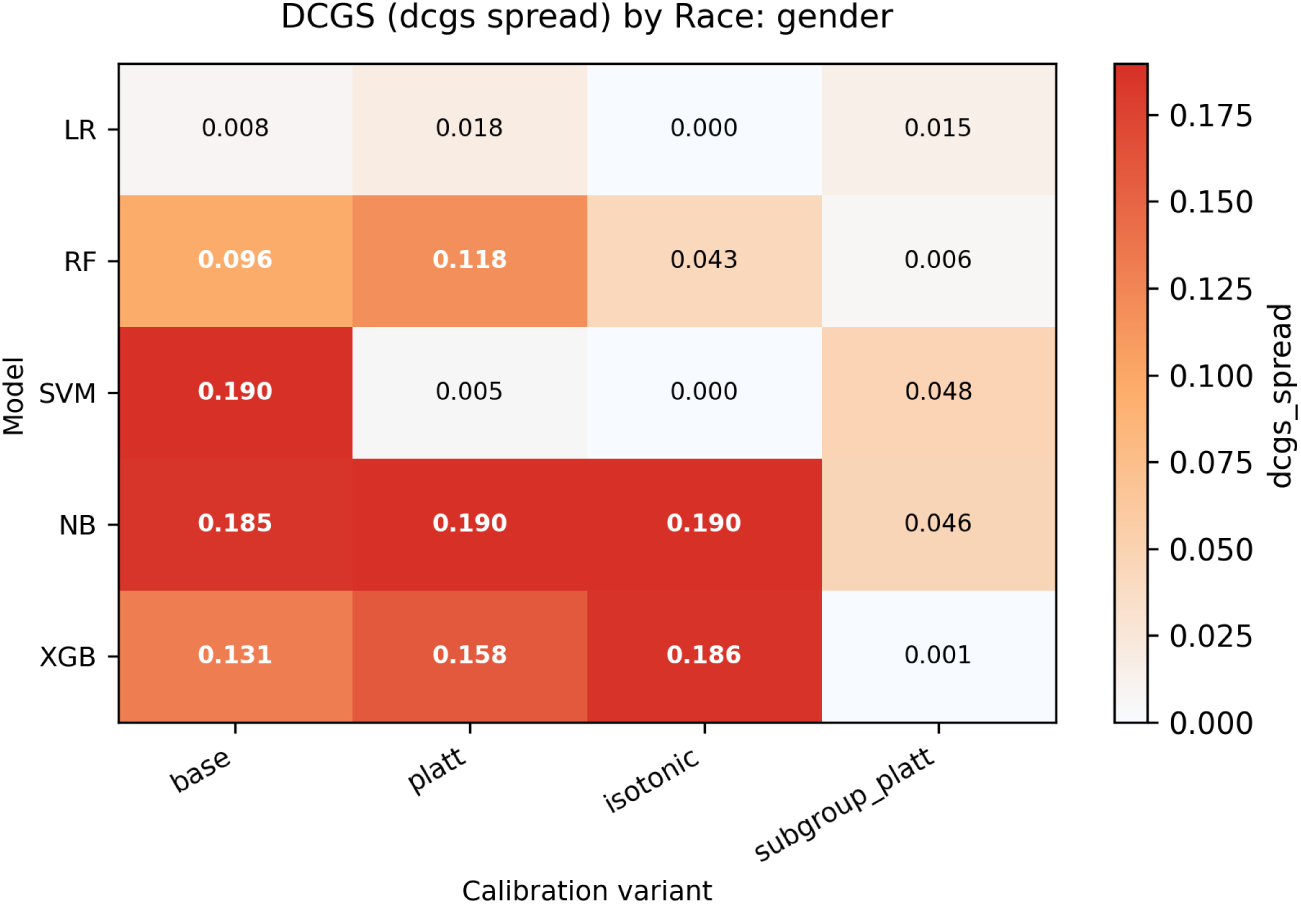
(b) DCGS spread heatmap by gender. Cell values show the standard deviation of ECE across gender subgroups.

**Table 5:** Conformal prediction coverage and Demographic Coverage Gap (MIMIC-IV evaluation half, n=658). Target coverage: 90% (alpha=0.10).

| Model | Calibration | Threshold | Wis. coverage | MIMIC coverage | DCG race | DCG gender |
| --- | --- | --- | --- | --- | --- | --- |
| LR | base | 1.000 | 0.439 | 0.263 | 0.156 | 0.335 |
| LR | platt | 0.997 | 0.474 | 0.252 | 0.195 | 0.364 |
| LR | isotonic | 1.000 | 0.368 | 0.249 | 0.195 | 0.373 |
| LR | subgroup_platt | 0.997 | 0.474 | 0.336 | 0.406 | 0.135 |
| RF | base | 1.000 | 0.368 | 0.249 | 0.195 | 0.373 |
| RF | platt | 0.978 | 0.368 | 0.249 | 0.195 | 0.373 |
| RF | isotonic | 1.000 | 0.368 | 0.249 | 0.195 | 0.373 |
| RF | subgroup_platt | 0.978 | 0.368 | 0.249 | 0.195 | 0.373 |
| SVM | base | 0.999 | 0.526 | 0.249 | 0.195 | 0.373 |
| SVM | platt | 0.982 | 0.509 | 0.258 | 0.156 | 0.348 |
| SVM | isotonic | 1.000 | 0.368 | 0.249 | 0.195 | 0.373 |
| SVM | subgroup_platt | 0.982 | 0.509 | 0.249 | 0.195 | 0.373 |
| NB | base | 1.000 | 0.368 | 0.249 | 0.195 | 0.373 |
| NB | platt | 0.950 | 0.509 | 0.249 | 0.195 | 0.373 |
| NB | isotonic | 1.000 | 0.368 | 0.249 | 0.195 | 0.373 |
| NB | subgroup_platt | 0.950 | 0.509 | 0.277 | 0.305 | 0.298 |
| XGB | base | 1.000 | 0.430 | 0.249 | 0.195 | 0.373 |
| XGB | platt | 0.961 | 0.456 | 0.283 | 0.101 | 0.334 |
| XGB | isotonic | 1.000 | 0.368 | 0.249 | 0.195 | 0.373 |
| XGB | subgroup_platt | 0.961 | 0.456 | 0.249 | 0.195 | 0.373 |

DCG: Demographic Coverage Gap = $\max(\text{coverage}_k) - \min(\text{coverage}_k)$ across subgroups with at least 30 members.

Subgroup-targeted Platt scaling addresses this directly by fitting separate calibration functions per group. RF subgroup_platt DCGS_race is 0.124, below its base (0.128) and Platt (0.122) versions. LR subgroup_platt achieved DCGS_race of 0.071, lower than its base (0.089). The reductions are real, but none bring DCGS below 0.05. Subgroup calibration samples on the MIMIC calibration half ranged from roughly 30 to 148 patients per racial group, too small for the Platt function to fit reliably.

The OTHER racial category was persistently the hardest to calibrate. It aggregates multiple identities that do not fit the five-group taxonomy used here. That internal heterogeneity prevents any single calibration function from working well across the full group. Collecting and reporting finer-grained demographic data in clinical AI datasets is a direct consequence.

### Clinical implications

The 0.05 DCGS threshold is not arbitrary. A model assigning a 30% probability of malignancy to a Hispanic woman should mean the same thing as a 30% probability assigned to a White woman. When DCGS is 0.19 on the race axis, those numbers mean different things for different groups. A biopsy referral threshold set at 25% then consistently places patients below or above the line for the wrong reasons.

The numbers put this in perspective. Approximately 39 million mammograms are performed annually in the United States [27]. At AI-assisted risk scoring penetration rates between 5% and 25%, a DCGS above 0.05 affects between 2 million and 10 million case evaluations per year. At 10% penetration the figure is approximately 4 million. Black and Hispanic women, already facing documented mortality disparities, carry the largest share of that error [3, 17].

Adding DCGS to standard evaluation workflows is straightforward. Compute ECE per subgroup. Take the range and standard deviation. Report them alongside the global ECE in the calibration table. The additional computation takes fewer than ten lines of code. Low barrier. The reason this has not been standard practice appears to be that no prior work named and formalized it.

The conformal coverage results tell a separate story. A 90% marginal coverage target produced actual MIMIC coverage near 25% for most model-variant combinations. That collapse was expected given the distributional mismatch. More telling: even within the MIMIC cohort, racial and gender coverage gaps exceeded 0.15 for all 20 combinations. Coverage already low in aggregate was not shared equally across groups. This pattern aligns with work from Zhou and Sesia and Lu et al. showing that split conformal prediction’s marginal guarantee does not extend to subgroups [24, 25]. Equalized coverage requires group-specific calibration thresholds, which in turn require larger calibration samples per demographic group than most clinical datasets currently provide.

### Limitations

The MIMIC cohort is from a single center. The demographic distribution and clinical practice patterns at Beth Israel Deaconess Medical Center do not represent the broader US hospital population. Racial subgroup sizes in the MIMIC evaluation half range from roughly 30 (ASIAN) to 150 (WHITE), making ECE estimates for smaller groups imprecise.

The feature mismatch between Wisconsin and MIMIC is severe by design. The near-chance AUROC on MIMIC says nothing about these models’ clinical utility. The relevant finding is that calibration gaps persist even when overall discrimination is poor, which is the condition most likely during deployment on out-of-distribution populations.

Race and gender are examined as single-axis subgroups here. Intersectional analysis requires sample sizes this cohort cannot support. Age tier and insurance subgroups available in the MIMIC-IV hospital module are absent from the ED module used here. Full MIMIC-IV hospital access would allow those axes to be included in future work.

## 5 Conclusion

Population-level calibration metrics are not enough for equitable clinical AI. A model with ECE of 0.02 on an internal test set produces DCGS above 0.19 on an external cohort. That gap is invisible without subgroup-stratified reporting.

DCGS makes these disparities visible. The 0.05 threshold gives a concrete criterion: when DCGS exceeds it, global recalibration is not enough and subgroup-targeted approaches are warranted. No single calibration method reliably brought DCGS below 0.05 across model types and subgroup axes in these experiments. Researchers should compute DCGS on their own external cohort rather than assuming any calibration method will close demographic gaps.

The DCGS metric, computation library, and full evaluation pipeline are available as open-source Python code under the MIT License at github.com/MichaelEnny/breast-cancer-calibration-gaps. The work is part of a broader research program on calibration equity in healthcare AI spanning multiple clinical domains and datasets.

## Acknowledgments

The author thanks the PhysioNet platform and Beth Israel Deaconess Medical Center for maintaining public access to the MIMIC-IV dataset. The Wisconsin Diagnostic Breast Cancer dataset was obtained from the UCI Machine Learning Repository. No external individuals contributed to the analysis.

## Author Contributions

Michael. O. Eniolade: Conceptualization, methodology, data collection, software development, formal analysis, writing, review, and editing.

## Ethical Approval

MIMIC-IV is a de-identified publicly available database. Access was obtained through PhysioNet following completion of the required data use agreement and CITI Program training. The Wisconsin Diagnostic Breast Cancer dataset is in the public domain. No primary patient data were collected for this study and institutional review board approval was not required.

## Data Availability

The Wisconsin Diagnostic Breast Cancer dataset is publicly available from the UCI Machine Learning Repository (dataset id=17). Access to MIMIC-IV requires credentialed PhysioNet registration. Code, the DCGS computation library, and all analysis scripts are available at github.com/MichaelEnny/breast-cancer-calibration-gaps under the MIT License.

## References

[1] S. Hussain et al., “Breast cancer risk prediction using machine learning: a systematic review,” Front. Oncol., vol. 14, p. 1343627, 2024, doi: 10.3389/fonc.2024.1343627.

[2] C. L. Andaur Navarro et al., “Completeness of reporting of clinical prediction models developed using supervised machine learning: a systematic review,” BMC Med. Res. Methodol., vol. 22, no. 1, p. 12, 2022.

[3] C. E. DeSantis et al., “Breast cancer statistics, 2019,” CA Cancer J. Clin., vol. 69, no. 6, pp. 438–451, 2019.

[4] T. Ganta et al., “Fairness in predicting cancer mortality across racial subgroups,” JAMA Netw. Open, vol. 7, no. 7, p. e2421290, 2024, doi: 10.1001/jamanetworkopen.2024.21290.

[5] M. Hardt, E. Price, and N. Srebro, “Equality of opportunity in supervised learning,” Adv. Neural Inf. Process. Syst., vol. 29, pp. 3315–3323, 2016.

[6] U. Hebert-Johnson, M. P. Kim, O. Reingold, and G. N. Rothblum, “Multicalibration: Calibration for the (computationally-identifiable) masses,” in Proc. 35th Int. Conf. Mach. Learn. (ICML), vol. 80, 2018, pp. 1939–1948.

[7] A. E. W. Johnson et al., “MIMIC-IV, a freely accessible electronic health record dataset,” Sci. Data, vol. 10, no. 1, p. 1, 2023.

[8] M. Gupta et al., “An extensive data processing pipeline for MIMIC-IV,” in Proc. 2nd Conf. Mach. Learn. Health (ML4H), PMLR, vol. 193, 2022, pp. 311–325, doi: 10.48550/arXiv.2204.13841.

[9] M. O. Eniolade, “Calibration, uncertainty communication, and deployment readiness in CKD risk prediction: A framework evaluation study,” arXiv preprint arXiv:2605.21566, 2026.

[10] B. Van Calster et al., “Calibration: the Achilles heel of predictive analytics,” BMC Med., vol. 17, no. 1, p. 230, 2019.

[11] Y. Huang, W. Li, F. Macheret, R. A. Gabriel, and I. Ohno-Machado, “A tutorial on calibration measurements and calibration models for clinical prediction models,” J. Am. Med. Inform. Assoc., vol. 27, no. 4, pp. 621–633, 2020.

[12] J. C. Platt, “Probabilistic outputs for support vector machines and comparisons to regularized likelihood methods,” in Adv. Large Margin Classifiers, A. Smola, P. Bartlett, B. Scholkopf, and D. Schuurmans, Eds. Cambridge, MA, USA: MIT Press, 1999, pp. 61–74.

[13] B. Zadrozny and C. Elkan, “Transforming classifier scores into accurate multiclass probability estimates,” in Proc. 8th ACM SIGKDD Int. Conf. Knowl. Discov. Data Min., 2002, pp. 694–699.

[14] W. G. La Cava, E. Lett, and G. Wan, “Fair admission risk prediction with proportional multicalibration,” in Proc. Conf. Health, Inference, Learn. (CHIL), pp. 350–378, 2023.

[15] C. Shui, J. Szeto, R. Mehta, D. L. Arnold, and T. Arbel, “Mitigating calibration bias without fixed attribute grouping for improved fairness in medical imaging analysis,” in Int. Conf. Med. Image Comput. Comput.-Assist. Intervent. (MICCAI), pp. 189–198, 2023.

[16] R. J. Chen et al., “Algorithmic fairness in artificial intelligence for medicine and healthcare,” Nat. Biomed. Eng., vol. 7, pp. 719–742, 2023.

[17] C. G. Yedjou et al., “Health and racial disparity in breast cancer,” Adv. Exp. Med. Biol., vol. 1152, pp. 31–49, 2019.

[18] R. B. Hines et al., “Health insurance and neighborhood poverty as mediators of racial disparities in advanced disease stage at diagnosis and nonreceipt of surgery for women with breast cancer,” Cancer Med., vol. 12, pp. 15414–15423, 2023, doi: 10.1002/cam4.6127.

[19] A. Soltan and P. Washington, “Challenges in reducing bias using post-processing fairness for breast cancer stage classification with deep learning,” Algorithms, vol. 17, no. 4, p. 141, 2024, doi: 10.3390/a17040141.

[20] R. B. Parikh, C. R. Manz, C. Chivers, et al., “Machine learning approaches to predict 6-month mortality among patients with cancer,” JAMA Netw. Open, vol. 2, no. 10, p. e1915997, 2019.

[21] I. Dankwa-Mullan and D. Weeraratne, “Artificial intelligence and machine learning technologies in cancer care: addressing disparities, bias, and data diversity,” Cancer Discov., vol. 12, no. 6, pp. 1423–1427, 2022, doi: 10.1158/2159-8290.CD-21-1601.

[22] Z. Obermeyer, B. Powers, C. Vogeli, and S. Mullainathan, “Dissecting racial bias in an algorithm used to manage the health of populations,” Science, vol. 366, no. 6464, pp. 447–453, 2019.

[23] A. N. Angelopoulos and S. Bates, “Conformal prediction: A gentle introduction,” Found. Trends Mach. Learn., vol. 16, no. 4, pp. 494–591, 2023, doi: 10.1561/2200000101.

[24] Y. Zhou and M. Sesia, “Conformal classification with equalized coverage for adaptively selected groups,” Adv. Neural Inf. Process. Syst., vol. 37, pp. 108760–108823, 2024.

[25] C. Lu, A. Lemay, K. Chang, K. Höbel, and J. Kalpathy-Cramer, “Fair conformal predictors for applications in medical imaging,” in Proc. AAAI Conf. Artif. Intell., vol. 36, no. 11, 2022, pp. 12008–12016, doi: 10.1609/aaai.v36i11.21459.

[26] W. N. Street, W. H. Wolberg, and O. L. Mangasarian, “Nuclear feature extraction for breast tumor diagnosis,” in Proc. SPIE Int. Symp. Electron. Imaging Sci. Technol., vol. 1905, 1993, pp. 861–870.

[27] B. F. Churchill and E. C. Lawler, “Government recommendations and health behaviors: Evidence from breast cancer screening guidelines,” Natl. Bur. Econ. Res. Working Paper 34368, 2025, doi: 10.3386/w34368.

[28] C. Guo, G. Pleiss, Y. Sun, and K. Q. Weinberger, “On calibration of modern neural networks,” in Proc. 34th Int. Conf. Mach. Learn. (ICML), vol. 70, 2017, pp. 1321–1330.

[29] M. P. Naeini, G. Cooper, and M. Hauskrecht, “Obtaining well calibrated probabilities using Bayesian binning,” in Proc. AAAI Conf. Artif. Intell., vol. 29, no. 1, pp. 2901–2907, 2015, doi: 10.1609/aaai.v29i1.9602.

[30] E. W. Steyerberg et al., “Assessing the performance of prediction models: a framework for traditional and novel measures,” Epidemiology, vol. 21, no. 1, pp. 128–138, 2010.

[31] S. Aamir et al., “Predicting breast cancer leveraging supervised machine learning techniques,” Comput. Math. Methods Med., vol. 2022, p. 5869529, 2022, doi: 10.1155/2022/5869529.

[32] H. Chen, N. Wang, D. Xie, K. Mei, Y. Zhou, and G. Cai, “Classification prediction of breast cancer based on machine learning,” Comput. Intell. Neurosci., vol. 2023, Art. no. 6530719, 2023, doi: 10.1155/2023/6530719.

[33] M. M. Islam et al., “Enhancing breast cancer detection and classification using advanced multi-model features and ensemble machine learning techniques,” Diagnostics, vol. 13, no. 19, p. 3106, 2023, doi: 10.3390/diagnostics13193106.

[34] M. M. Obare, “Survey and comparative analysis of machine learning algorithms for breast cancer diagnosis: A comprehensive review,” World J. Adv. Res. Rev., vol. 19, no. 1, pp. 1136–1149, 2023, doi: 10.30574/wjarr.2023.19.1.1464.

[35] C. L. Andaur Navarro, J. A. A. Damen, M. van Smeden, T. Takada, S. W. J. Nijman, P. Dhiman, J. Ma, G. S. Collins, R. Bajpai, R. D. Riley, K. G. M. Moons, and L. Hooft, “Systematic review identifies the design and methodological conduct of studies on machine learning-based prediction models,” J. Clin. Epidemiol., vol. 154, pp. 8–22, 2023, doi: 10.1016/j.jclinepi.2022.11.015.

